# An altered B cell compartment distinguishes low antibody responders among people with HIV

**DOI:** 10.64898/2026.09.18.26363407

**Authors:** Karli R. Redinger, Samuel B. Warner, Armaan Jamal, Nuria Gallego Marquez, Matthew L. Mendoza, David E. Sanin, Michael J. Peluso, Alan L. Landay, Andrea L. Cox, Annukka A. R. Antar, Elizabeth A. Thompson

## Abstract

Chronic HIV infection can leave persistent immune dysregulation despite effective antiretroviral therapy (ART), but how this altered immune landscape affects antibody induction and durability after antigen exposure remains unclear. We used SARS-CoV-2 infection and vaccination as a model of humoral immunity to a novel antigen, evaluating anti-RBD IgG titers and immune cell phenotypes in people with HIV (PWH) and without HIV (PWoH) with hybrid immunity, stratified by early (<90 days) or late (≥90 days) post-exposure time points. PWH who failed to mount protective anti-RBD IgG titers early had a B cell compartment that was distinct from PWoH and high-titer PWH. Specifically, low-titer PWH had the highest expansion of activated naïve, CD11c⁺ age-associated, and IgM-skewed B cells with reduced regulatory marker expression, even after controlling for age, sex, BMI and days since infection. PWH who had low-titer responses during the late phase of the response had persistent CD11c-associated features with expansion of double-negative and altered class-switched memory B cell compartments. These B cell features correlated with CD4 and CD8 T cell activation and senescence-associated phenotypes, suggesting that failure to mount high titer antibody response and maintenance in a subset of PWH is shaped by coordinated immune dysregulation.

## Introduction

The COVID-19 pandemic has caused immense global suffering, resulting in millions of deaths and widespread hardship. However, at the same time, the pandemic and subsequent vaccination campaign offered an unprecedented opportunity to study infection-and vaccine-induced immune responses to a novel antigen at the population level. Since the onset of the pandemic, there have been approximately 779 million cases and over 13.6 billion vaccines rolled out globally^1,2^. At this point in time, almost everyone has been exposed through vaccination and infection thus allowing for the exploration of immune responses among different populations, potentially revealing insights about how people mount productive responses to inform future vaccine design. Such insights may ultimately help prevent another large-scale global health crisis of this magnitude.

SARS-CoV-2 humoral responses have been very well characterized in healthy people, with both infection and vaccination inducing robust neutralizing antibody (Ab) titers that peak within the first three weeks and begin to wane within six to eight months post-exposure^3–5^. However, various immunocompromised and immune dysregulated populations have lower humoral responses following infection and vaccination and thus rely on successful vaccination for sufficient protection against severe disease. We have previously found that successful vaccination among solid organ transplant recipients relied more heavily on the expansion of CD11c+ B cells compared to healthy individuals^6^. Other groups have found that multiple immune dysregulated populations demonstrate consistent associations between poor vaccine responsiveness and the expansion of exhausted CD21low or CD27-IgD-double-negative B cells^7–10^. Conversely, successful protective immunity across these immunosuppressed cohorts was associated with higher frequencies of naïve B cells at baseline^11,12^. Together, these data indicate that alternative mechanisms for successful responses can exist in immune dysregulated populations.

In the present study, we investigated people with HIV (PWH), due to evidence of ongoing immune dysregulation despite successful HIV suppression with anti-retroviral therapy (ART). ART has drastically improved outcomes for PWH by restoring CD4 T cell counts, thereby mitigating the risk of severe infections, morbidity, and mortality when used as prescribed^13,14^. However, despite effective ART, PWH still experience varying levels of immune dysregulation thought to be due to systemic inflammation, mucosal damage, microbiome disruption, lymphoid tissue fibrosis, and T cell exhaustion that accompany chronic viral infection^15–17^. As a result, studies have shown that, on average, PWH have lower responses to common vaccines including influenza and Hepatitis A/B compared to people without HIV^18–22^. More recently, the risk of adverse SARS-CoV-2 outcomes among PWH has been variable, with some studies showing PWH are more susceptible to severe infection while others show no difference between PWH and people without HIV^16,23–25^. Higher CD4 counts and well-controlled viremia are generally associated with improved vaccine responses and reduced disease severity^16,26–28^, while additional vaccine doses can increase seroconversion and partially overcome accelerated Ab decay^16,29–31^. These studies emphasize the heterogeneity of this population, which could be influenced by stage of HIV infection, time from HIV infection to initiation of ART, nadir and current CD4+ counts, and the extent of immune dysregulation despite ART as outlined above.

In addition to changes in T cell phenotype and activation during HIV infection, it is well established that HIV remodels the peripheral B cell compartment, which likely also contributes to humoral responses to vaccination and infection. PWH show persistent losses of resting and switched memory cells and expansion of transitional, activated-memory, tissue-like memory, and double-negative populations, and this dysregulation is only partly reversed by ART even with viral suppression^32–38^. However, how these alterations translate into changes in antibody responses remains less well characterized, and recent studies during the COVID-19 pandemic have begun to address this question, yet have primarily focused on major B cell subsets rather than deeper functional characterization^10,39,40^.

To this end, we investigated SARS-CoV-2 humoral responses among PWH and asked which B cell profiles were associated with the induction and maintenance of SARS-CoV-2 Ab titers. To address these questions, we obtained SARS-CoV-2 antibody titers and performed high parameter deep profiling of the B cell compartment in a subset of PWH and people without HIV (PWoH) who had hybrid SARS-CoV-2 immunity from a larger JHU-HIV-SARS-CoV-2 cohort. Consistent with prior literature, we found that PWH exhibit alterations in the B cell compartment overall, but among those with low antibody titers, these changes were marked by enrichment of CD11c⁺ activated naïve and age-associated B cells, along with other atypical B cell phenotypes. These low-titer-associated features also correlated with T cell activation, supporting a model in which a subset of PWH has coordinated immune dysregulation linked to poor antibody responses.

## Results

### Overview of clinical cohort and sample selection

In the JHU-HIV-SARS-CoV-2 cohort, participants were recruited between June 2021 and February 2023. The cohort was designed to prospectively assess long-term symptoms, post-acute sequelae, and cognitive outcomes 12 months after SARS-CoV-2 infection in people with and without HIV. Participants were enrolled into four study arms, including individuals with prior SARS-CoV-2 infection and uninfected controls, with and without HIV: (1) PWH who had their first bout of COVID-19 within the previous 4 weeks, (2) HIV negative individuals who had their first bout of COVID-19 within the previous 4 weeks, (3) PWH who had never had COVID-19 to their knowledge, and (4) HIV negative individuals who had never had COVID-19^41^. In total, 150 PWH and 182 PWoH were recruited (**Figure S1)**. Blood was collected at approximately 1 and 4 months post-COVID symptom onset for individuals recruited with recent COVID-19, or collected one time for individuals who had never experienced COVID-19. The study collected demographic and clinical information such as last reported HIV viral load, last reported CD4 count, and days since last SARS-CoV-2 infection or vaccination. The study also quantified CD4 count from freshly processed blood samples for PWH. To quantify humoral responses after SARS-CoV-2 antigen exposure, the parent cohort measured ancestral anti-spike and anti-RBD IgG titers in all participants.

For the present sub-analysis correlating B cell phenotypes with induction of humoral immunity, we performed deep immune profiling using spectral flow cytometry in 155 samples from the parent cohort (**Figure S1**) and selected 123 samples (44 from PWH and 79 from PWoH) from participants with hybrid immunity, defined as having had both prior SARS-CoV-2 infection and vaccination prior to sampling. This restriction was applied to minimize heterogeneity in infection and vaccination histories and to focus on individuals expected to have measurable anti–SARS-CoV-2 Ab responses, enabling assessment of immune features associated with response. Individuals without prior SARS-CoV-2 infection or without vaccination were excluded for subsequent analysis, unless used to compare B cell frequencies to account for differences due to recent infection.

To finalize sample selection and categorization, we first stratified participants by time since most recent antigen exposure into an early post-exposure interval, defined as <90 days since antigen exposure, and a late post-exposure window, based on previous studies indicating that protective Ab titers rapidly decline within the first three months post-exposure^42–44^ **(Figure 1A, vertical line)**. Consistent with the literature, anti-spike and anti-RBD IgG titers waned over time for both PWoH and PWH, regardless of whether the last antigen exposure was infection or vaccination **(Figure 1A and S2A-C)**. Because some individuals received an additional vaccination between study visits, samples were analyzed by post-exposure interval rather than as a matched longitudinal series. When repeat samples from the same individual fell within the same <90-day interval after antigen exposure, one sample was excluded to avoid overrepresentation. The final samples in the subsequent analysis included 105 samples (39 from PWH and 66 from PWoH, **Figure S1**). In this subsampling of participants, PWH and PWoH were matched on all variables (age, BMI, last antigen exposure type, and number of vaccine doses) with the exception of sex **(Table S1)**, reflecting the unbalanced sex recruitment in the overall cohort. All but one individual with HIV included in this sub study reported that their last HIV viral load was undetectable.

**Figure 1:**
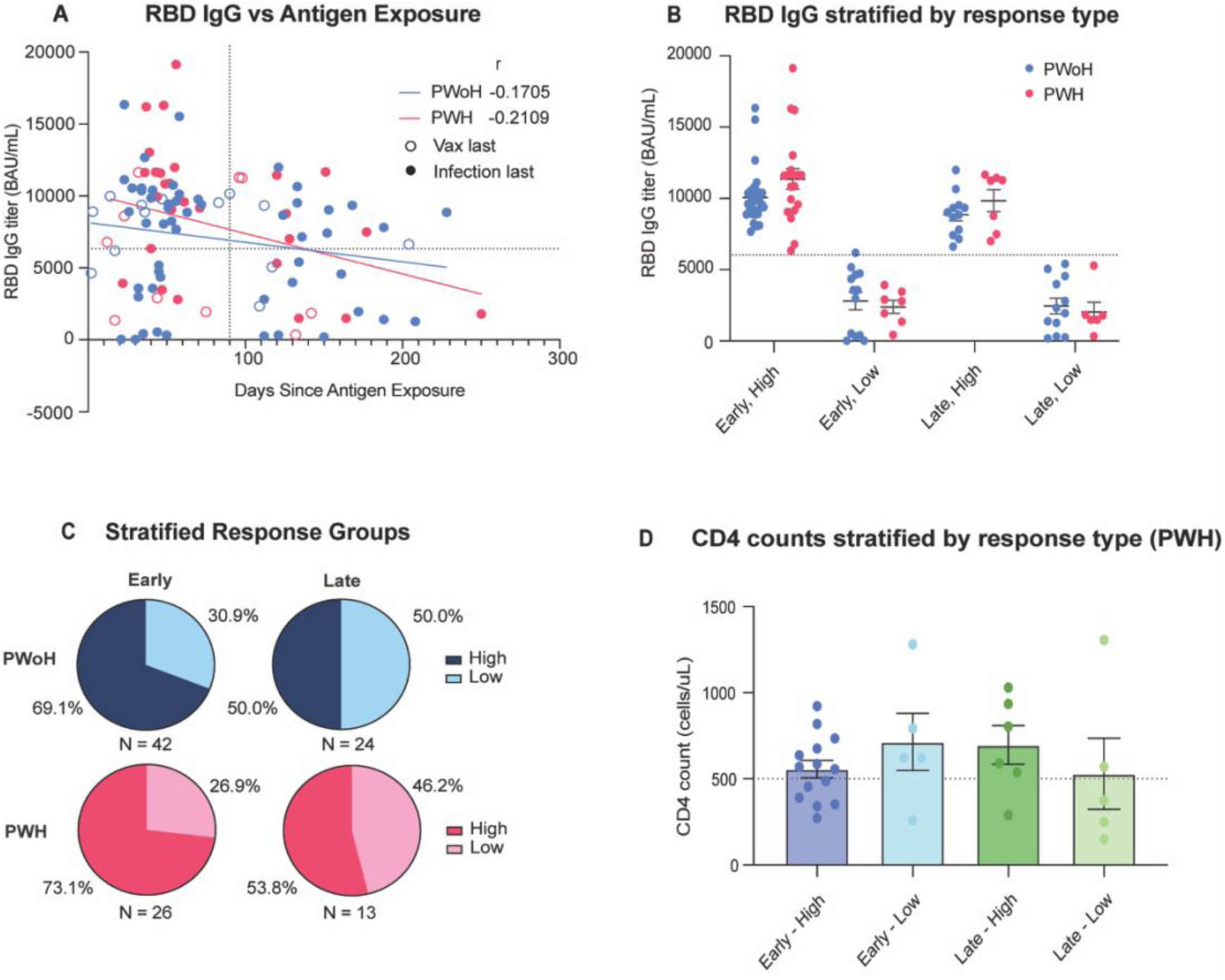
SARS-CoV-2 antibody responses do not correlate with common clinical factors for PWH. **A.** Anti-RBD IgG titers plotted against days since most recent antigen exposure for PWoH (n = 66) and PWH (n = 39). Horizontal dotted line denotes titer threshold for high versus low titer individuals (6321 BAU/mL). Vertical dotted line denotes cutoff for early versus late time point (90 days). **B**. Anti-RBD IgG titers for PWoH and PWH stratified by Ab titer status. Significance tested using Mann-Whitney U test. **C.** Proportion of PWoH and PWH with high vs low titers stratified by response time. **D.** Anti-RBD IgG titers stratified by Ab titer status for PWH. Horizontal line denotes the clinical threshold for high versus low CD4 count (500 cell/uL). Significance tested using Kruskal-Wallis test. Data presented as Mean±SEM. All correlations were performed using Spearman correlation.

We further designated PWoH and PWH as either high or low Ab titer responders using a previously reported protective threshold against infection acquisition for anti-RBD IgG based on a recent household contact study (6321 BAU/mL)^45^ **(Figure 1A, horizontal line)**. Although this cutoff is not a universal correlate of protection and may vary by viral variant and exposure history, it provided a consistent cutoff for defining high-vs low-titer individuals. After stratification by time since antigen exposure, PWoH and PWH had similar anti-RBD IgG titers within each group **(Figure 1B)**, and low titer individuals represented a greater proportion of both groups in the later time points **(Figure 1C).** For PWH, each Ab response group contained individuals with both high and low clinical CD4 counts (available for 30/39 samples) **(Figure 1D)**, emphasizing that CD4 counts alone do not explain differences in Ab titer induction or maintenance. Of note, the one individual with a detectable self-reported viral load induced and maintained high Ab titers both early and late in the response.

### A distinct total B cell compartment characterized by CD11c expression and lack of class-switched memory distinguishes PWH who fail to mount robust early Ab titers

To identify alterations in the B cell compartment that could influence the induction and maintenance of Ab titers, PBMCs from all selected participants were analyzed by flow cytometry to evaluate differences in the induction of spike-specific B cells and the overall composition of B cell subsets. We and others have recently shown the importance of CD11c+ B cells in supporting vaccine responses in both healthy and immunosuppressed populations^6,46,47^. Therefore, we gated on canonical B cell subsets^48,49^, with an added focus on CD11c+ B cell subsets (**Figure S3A**). **Figure 2A** shows the average relative distribution of all B cell subsets analyzed across groups, including both significant and non-significant comparisons, and demonstrates a unique B cell landscape specifically in low-titer PWH compared to the other three groups. We then examined individual subset frequencies in greater detail. In this analysis, low-titer PWH exhibited higher frequencies of multiple canonical B cell subsets compared with the other three groups, although several pairwise comparisons were not significant and are shown in **Supplementary Figure S3B**. Among antigen-specific populations, PWoH demonstrated significantly higher frequencies of spike-specific and CD11c+ spike-specific B cells in high responders, consistent with their higher Ab titers, whereas no significant differences in spike-specific B cell frequencies were observed between high and low responders among PWH (**Figure 2B**). These data suggest that lower Ab titers early after antigen exposure in PWH reflect B cell quality rather than quantity, with the most pronounced compartmental remodeling observed in low-titer PWH.

**Figure 2.**
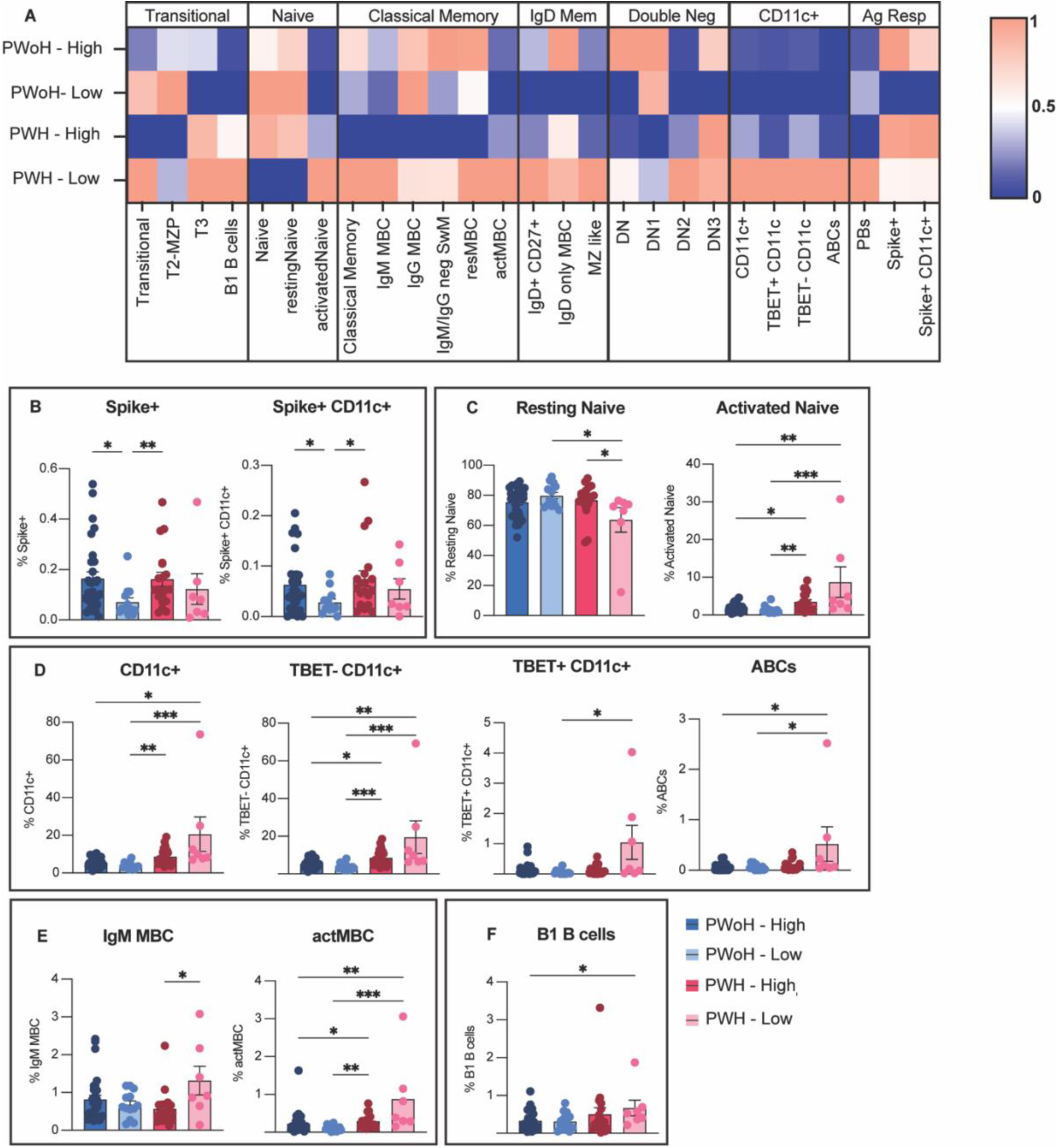
Low-titer PWH <G0 days post-antigen challenge have a distinct total B cell phenotype marked by CD11c expansion and lack of class-switched memory. **A.** Heatmap of normalized average frequencies of B cell subsets for PWoH and PWH stratified by Ab titer group. **B-F.** Frequencies of significantly different B cell subsets stratified by HIV status and Ab titer group. All frequencies are reported as percent of total B cells. Data presented as Mean±SEM. Significance tested using Kruskal-Wallis test. *p < 0.05, **p < 0.01, ***p < 0.001, and ****p < 0.0001

Consistent with potential defects in B cell quality, low-titer PWH sampled within 90 days post-antigen exposure showed enrichment of multiple B cell subsets marked by activation, CD11c expression, and IgM skewing (**Figure 2C-E)**. Within the naïve compartment, low-titer PWH had fewer resting naïve B cells, while PWH overall had increased activated naïve B cells, a subset defined in part by CD11c expression **(Figure 2C, Figure S3A)**. Direct analysis of CD11c further showed that low-titer PWH had significantly higher frequencies of total CD11c+ B cells compared to PWoH and a trend toward higher frequencies than high-titer PWH **(Figure 2D).** This CD11c+ enrichment in low-titer PWH included both TBET+ and TBET-CD11c+ B cells as well as age-associated B cells (ABCs, T-bet+/CD21lo/-/CD11c+), which have been associated with poor responses^50–52^. These data indicate that although CD11c expansion is on average higher in PWH compared to PWoH, higher levels of CD11c+ cells could be a defining feature of poor responses and may segregate high and low responders among PWH. In addition to their unique CD11c expression signature, PWH also had increased activated memory B cells (actMBC) compared to PWoH. In addition, although PWH broadly had increased activated memory B cells compared with PWoH, low-titer PWH were distinguished by higher frequencies of IgM⁺ memory B cells **(Figure 2E)** and B1-like B cells, which traditionally secrete IgM, compared to high-titer PWH (**Figure 2F**). These results may indicate defects in class switching or B–T cell interactions early in the response. In contrast, we found no significant differences among the four groups in other class-switched memory or early maturation compartments **(Figure S3B).** Importantly, key B cell subset differences identified in the primary unadjusted analyses remained significant after adjustment for age, sex, BMI, and days since symptom onset, suggesting that these covariates do not fully explain the observed B cell remodeling (**Figure S4**). Furthermore, we found no significant differences between these key subsets when stratifying by last antigen exposure type among PWH, and very few differences when comparing frequencies to COVID-controls indicating that these frequencies are not driven by recent infection **(Figure S5).** Given the differences in the frequencies of canonical B cell subsets, we next used high dimensional approaches to integrate B cell phenotypes and clinical factors together to determine which variables are most important in defining these different groups.

### Low-titer PWH early post-antigen exposure have decreased inhibitory marker expression and global B cell activation marked by CD11c+ and IgM+ expansion

Linear discriminant analysis (LDA) was performed using the pre-designated Ab titer groups to incorporate phenotypic marker expression in addition to B cell subset frequencies and clinical parameters to determine which variables (n = 363) (**Table S2**) contributed to group segregation. Given the high dimensionality of the dataset relative to cohort size, LDA was used as an exploratory data-reduction approach to identify candidate immunological and clinical features associated with titer group separation, rather than as a validated predictive classifier. This approach also allowed us to assess whether conventional clinical parameters or immunological features contributed more strongly to group discrimination. In line with our B cell frequency data in the early response, PWH low responders had the greatest segregation following LDA **(Figure 3A)**. We then employed an effect size filtering approach to identify 15 top variables (Cohen’s *d* > 1.0) important for group discrimination **(Figure S6)** followed by hierarchical clustering for visualization **(Figure 3B).** Of note, none of the clinical variables (including recent infection) were identified as top variables after effect size filtering, indicating that immunological profiles induced in a subset of PWH are most important when determining differences between the groups during the early response. In order to better characterize these phenotypic differences and identify functionally related variables, we calculated pairwise Pearson correlations among the top identified variables followed by hierarchical clustering to establish 5 distinct clusters (**Figure 3C).** Composite scores were subsequently generated for each cluster by calculating averaged z-scores, and each cluster was manually annotated based on biological variables to facilitate interpretation **(Figure 3D).**

**Figure 3.**
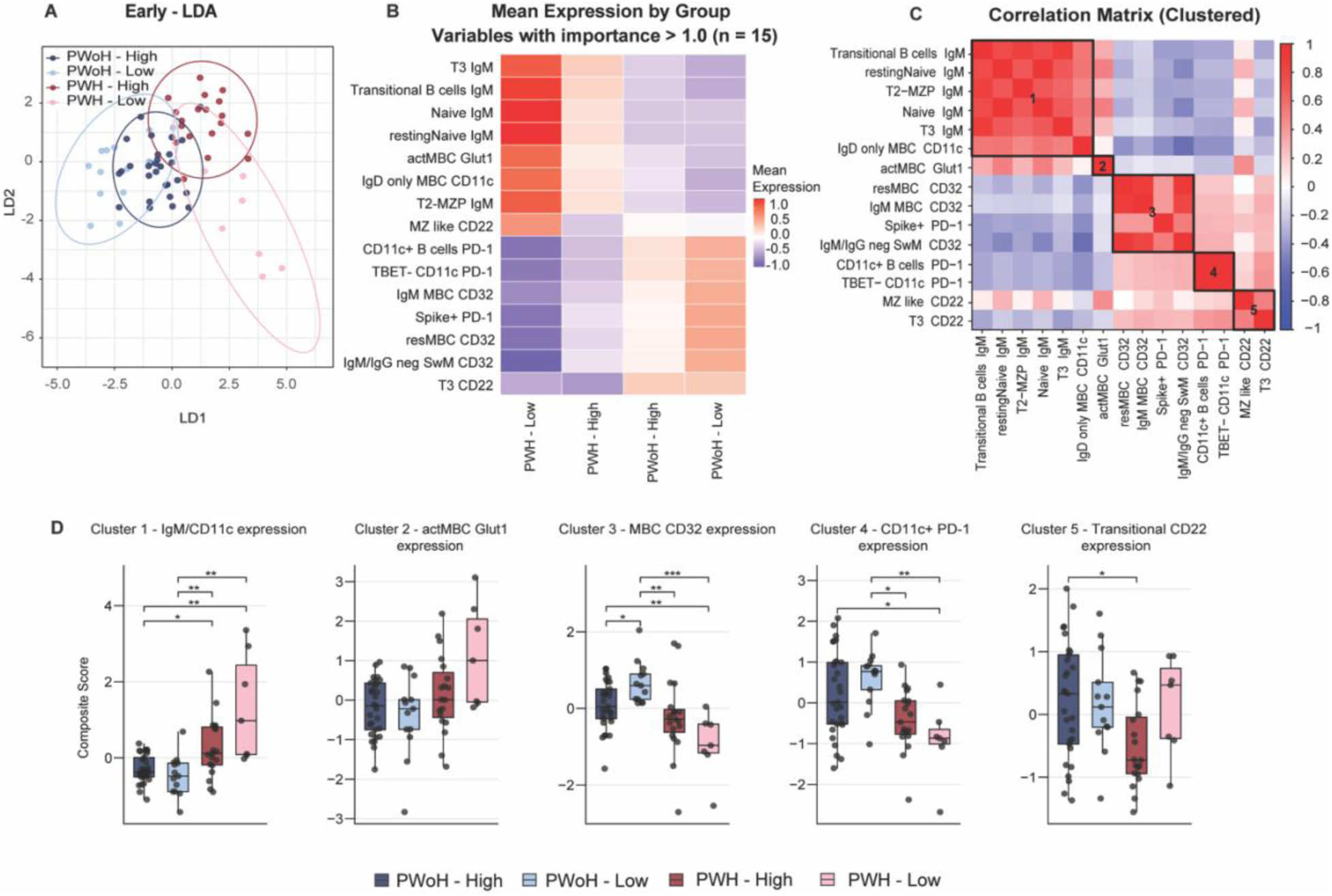
Low-titer PWH early-post antigen challenge have higher IgM, CD11c and decreased PD-1, CD32 expression on B cells. **A.** Linear discriminant analysis (LDA) projection based on 363 B cell phenotypic and clinical variables. Ellipses represent 95% confidence for each group. **B.** Heatmap of mean variable expression of type top variables stratified by Ab titer status with hierarchical clustering by group and variables. Top variables were determined using Cohen’s d effect size (d > 1.0) **C.** Spearman correlation matrix of top variables. Clusters were designated using complete linkage with a dendrogram cutoff at 0.7. **D.** Composite score distributions generated using averaged z-scores for manually annotated clusters. Significance tested using Kruskal-Wallis test. *p < 0.05, **p < 0.01, ***p < 0.001, and ****p < 0.0001

Comparison of composite scores across titer groups identified several B cell profiles that distinguished PWH from PWoH and suggested features associated with high-or low-titer status among PWH. Interestingly, CD32 (FcyRII) and PD-1 expression were lower across multiple B cell subsets in PWH compared with PWoH, with the lowest average expression observed in low-titer PWH, although differences between PWH groups did not reach significance (**Figure 3D**, cluster 3,4). While CD32 has both activation (CD32a) and inhibitory (CD32b) isoforms, CD32b is the predominant isoform expressed on human B cells^53–55^. Reduced CD32b expression has been associated with chronic B cell activation in autoimmune settings^56–59^, while PD-1 can mark recently activated post-germinal center B cells^60^ suggesting that lower expression of these markers may reflect a less regulated or less productively differentiated B cell state. Consistent with this interpretation, low-titer PWH also showed higher IgM expression across multiple B cell subsets, which correlated with CD11c expression (**Figure 3C–D**, cluster 1), potentially indicating skewing toward activated, unswitched, or extrafollicular-like B cell phenotypes. In contrast, high-titer PWH showed reduced CD22 expression on marginal zone (MZ)-like and transitional T3 B cells (**Figure 3D**, cluster 5). MZ-like B cells are innate-like polyreactive B cells^61–63^ and T3 B cells are often considered anergic autoreactive B cells^64–66^. Because CD22 raises the threshold for BCR activation^67,68^, lower CD22 expression on these noncanonical populations may suggest increased availability or activation potential of alternative B cell pools. Together, these exploratory analyses suggest that low-titer PWH are characterized by an IgM⁺CD11c⁺ B cell profile, whereas high-titer PWH may engage distinct noncanonical B cell pathways to support Ab production within an altered immune landscape.

### Failure to maintain protective Ab titers is associated with a distinct total B cell phenotype defined by atypical B cell expansion and an altered memory compartment in PWH

To identify features associated with Ab maintenance, we employed the same analysis approaches for samples collected ≥90d since antigen exposure. Low-titer PWH again displayed a distinct relative distribution of B cell subset frequencies compared with high-titer PWH and PWoH (**Figure 4A**). High-titer PWoH had increased frequencies of spike⁺CD11c⁺ B cells compared with low-titer PWoH, whereas these frequencies did not differ by titer status among PWH (**Figure 4B**). Total spike⁺ B cells showed similar but nonsignificant trends (**Figure S7**), suggesting that antigen-specific B cell frequency alone does not explain Ab titer status in PWH at later post-exposure intervals. Again, key B cell subset differences identified in the primary unadjusted analyses remained significant after adjustment for age, sex, BMI, and days since symptom onset (**Figure S8**).

**Figure 4.**
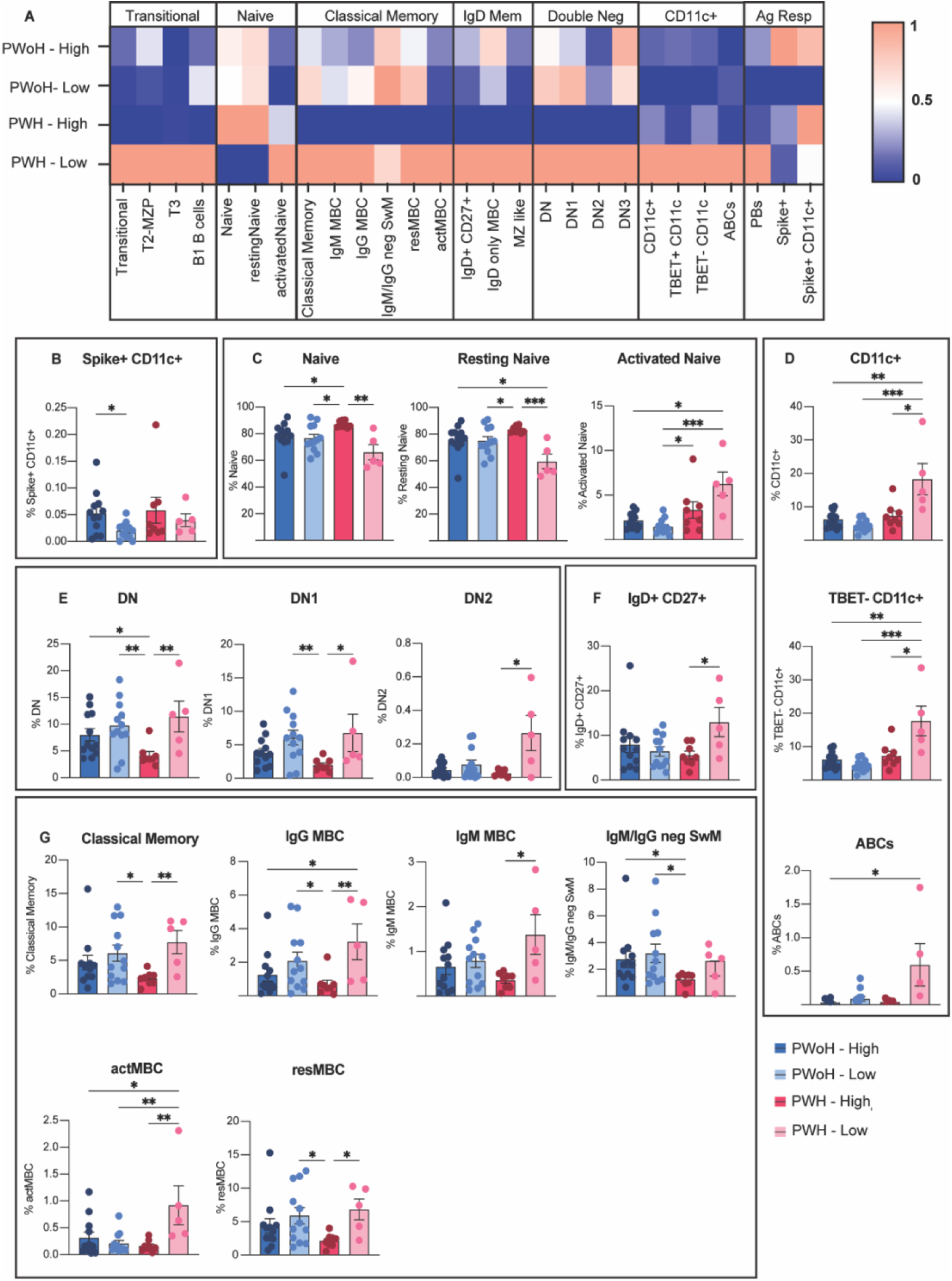
Low-titer PWH that fail to maintain protective Ab levels have expanded CD11c and atypical memory B cells. **A.** Heatmap of normalized average frequencies of B cell subsets for PWoH and PWH stratified by Ab titer status. **B-G.** Frequencies of significantly different B cell subsets stratified by HIV status and Ab titer status. All frequencies are reported as percent of total B cells. Data presented as Mean±SEM. Significance tested using Kruskal-Wallis test. *p < 0.05, **p < 0.01, ***p < 0.001, and ****p < 0.0001.

In line with earlier data, we found that low-titer PWH had increased activated naïve, CD11c+, and age-associated B cell frequencies **(Figure 4C-D)**. Consistent with this phenotype, low-titer PWH also had increased frequencies of double negative (IgD-CD27-) B cells compared to their high-titer counterparts, including CD11c+ DN2 cells **(Figure 4E)**. Because DN and CD11c⁺ B cell populations have been associated with aging, autoimmunity, and chronic immune activation^69–73^, their enrichment may reflect persistent inflammatory remodeling of the B cell compartment in low-titer PWH. These data suggest that cellular factors contributing to early vaccine failure are largely conserved late into the response, likely because these individuals lacked sufficient Ab titers from the outset.

However, there were also several unique cellular subsets identified that may independently drive long-term failure or success specifically in PWH, particularly with respect to differences in the memory compartment. Surprisingly, high-titer PWH had lower frequencies of multiple memory subsets including IgD⁺, IgM⁺, IgG⁺, and total classical memory B cell frequencies (**Figure 4F–G**). High-titer PWH also had lower frequencies of IgM⁻IgG⁻ class-switched memory B cells, potentially including IgA⁺ memory cells, although this subset requires further phenotypic confirmation. In contrast, high-titer PWH had increased frequencies of total and resting naïve B cells (**Figure 4C),** potentially reflecting a larger naïve repertoire to drive effective responses as demonstrated previously^11,12^. Together, these findings suggest that low Ab titers ≥90 days post-exposure in PWH are associated with persistent CD11c⁺/ABC enrichment and altered memory B cell differentiation rather than reduced spike-specific B cell frequency alone. Early B cell maturation subsets did not differ significantly among groups (**Figure S7**).

### Linear discriminant analysis identifies multiple biological signatures associated with low Ab titers among PWH later in the response

To determine if there were phenotypic differences in B cells that could impact Ab maintenance, we again performed LDA followed by hierarchal clustering utilizing the same variables as in the early time point **(Table S2)**. At this later time point, high-and low-titer PWH segregated into two distinct groups while PWoH largely overlapped **(Figure 5A)**. When performing effect size analysis of top variables, we found a total of 53 variables with an importance score greater than 1.0 **(Figure 5B, Figure S9)** indicating greater diversity in B cell phenotypes among the groups ≥90 days post-antigen challenge. However, we found that CD11c and IgM expression were again defining features among low-titer PWH in addition to the DN and classical memory frequencies identified previously **(Figure 5B)**.

**Figure 5.**
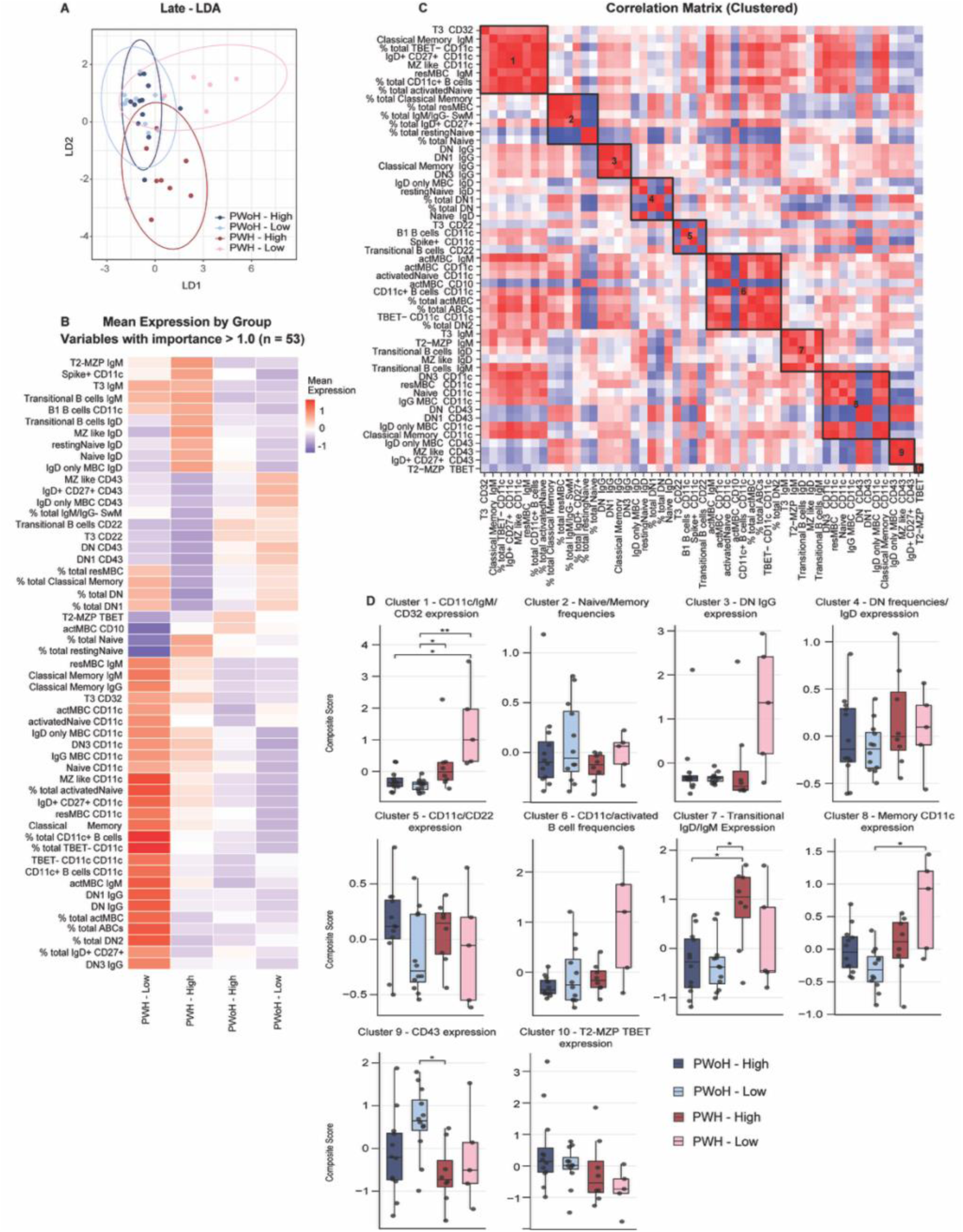
Multiple biological signatures define failure to maintain Ab titers in PWH. **A.** Linear discriminant analysis (LDA) projection based on 363 B cell phenotypic and clinical variables. Ellipses represent 95% confidence for each group. **B.** Heatmap of mean variable expression of type top variables stratified by response type with hierarchical clustering by group and variables. Top variables were determined using Cohen’s d effect size (d > 1.0) **C.** Spearman correlation matrix of top variables. Clusters were designated using complete linkage with a dendrogram cutoff at 0.7. **D.** Composite score distributions generated by PCA for manually annotated clusters. Significance tested using Kruskal-Wallis test. *p < 0.05, **p < 0.01, ***p < 0.001, and ****p < 0.0001.

In the same way clustering and composite scores were determined in the early time point, we identified and manually annotated 10 clusters of biologically related top variables among the late time point **(Figure 5C)**; however, many late clusters demonstrated high cellular diversity without reaching statistical significance among the Ab titer groups **(Figure 5D)**. This broader distribution of late-associated variables suggests that low Ab titers ≥90 days post-exposure may arise from more diverse B cell phenotypes than those observed within 90 days post-exposure, yet also may be reflective of lower sample size during the late time point. Despite this heterogeneity, certain profiles identified in the early time point were conserved in low-titer PWH over time, specifically characterized by CD11c expansion, IgM expression, and antigen-independent activation (**Figure 5D**, clusters 1, 6, 8). These data suggest that the profiles of early low-responders remain unchanged late in the response simply because there was no initial Ab response to maintain. In contrast, individuals who mounted robust initial Ab titers but lost them over time likely did so through multiple, distinct pathways, driving the high phenotypic diversity seen in the blood. Among high-titer PWH, we found greater expression of IgD and IgM on multiple transitional and naïve B cell subsets (**Figure 5D**, cluster 7). Higher BCR density on the surface of these subsets may allow for a lower threshold of activation upon stimulation and continual recruitment to ongoing GCs^74–77^, supporting our earlier findings that the transitional B cell pool may be utilized among PWH high responders to overcome chronic B cell exhaustion. Low-titer PWoH were distinguished by increased CD43 expression on IgD-expressing and MZ-like B cells. Because CD43 is largely absent from resting B cells and is upregulated upon activation, this pattern may reflect increased activation within an IgD⁺, innate-like B cell compartment rather than expansion of conventional class-switched memory responses^78,79^.

### PWH low responders have an inflammatory T cell profile marked by chronic activation

Because PWH may exhibit persistent T cell activation despite effective ART, which is often greatest among individuals with incomplete CD4 T cell reconstitution^17,80–82^, and because CD4 T cells are critical for B cell affinity maturation and class-switching, we next investigated whether peripheral T cell phenotypes correlated with the observed alterations in B cell phenotypes. Using a broad immune cell profiling flow cytometry panel, we extracted frequencies of CD4 and CD8 memory and naïve subsets as well as MFIs for key activation and differentiation markers on bulk CD4 and CD8 T cells **(Figure S10, Table S3).** We then performed Spearman correlations between these normalized T cell variables (n=31) and significant B cell composite scores independently for the early and late time points to identify coordinated cellular networks associated with high or low Ab titer groups (**Figure 6)**.

**Figure 6.**
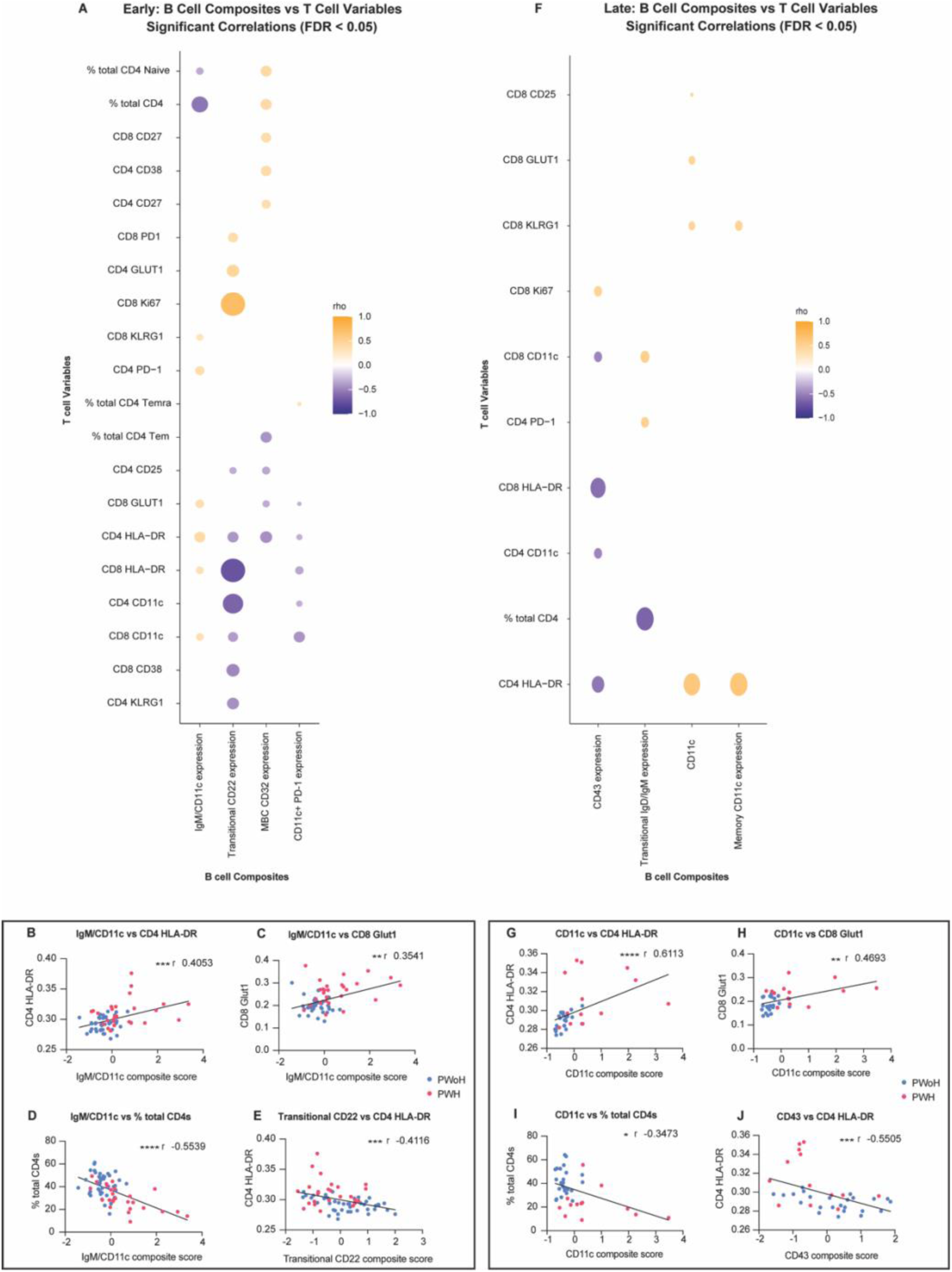
Inflammatory T cell phenotypes correlate with significant B cell clusters that define poor induction and maintenance of Ab titers. **A.** Correlation coefficient matrix of significant B cell clusters and T cell variables for the early time point. Significance was tested using Spearman rank correlations, and p-values were adjusted using false discovery rate (FDR < 0.05). Dot color represents correlation coefficients (ρ), and dot size represents statistical significance (-log₁₀(adjusted p-value)). **B-E**. T cell activation marker MFIs or % total CD4s plotted against respective B cell composite scores for all samples in the early time point. Pink dots denote PWH and blue dots denote PWoH. **F.** Correlation coefficient matrix of significant B cell clusters and T cell variables for the late time point. **G-J.** T cell activation marker MFIs or % total CD4s plotted against respective B cell composite scores for all samples in the late time point. Pink dots denote PWH and blue dots denote PWoH. All correlations were performed using Spearman correlation. *p < 0.05, **p < 0.01, ***p < 0.001, and ****p < 0.0001

Within 90 days post-exposure, B cell features enriched in low-titer PWH were associated with systemic T cell activation (**Figure 6A**). Increased B cell IgM/CD11c expression positively correlated with both CD4 and CD8 T cell activation markers, including CD4 HLA-DR **(Figure 6B)** and CD8 Glut1 **(Figure 6C)**, whereas B cell PD-1 expression was inversely associated with this profile. These T cell activation features were negatively correlated with total CD4 T cell frequencies **(Figure 6D)**, consistent with a relationship between incomplete CD4⁺ T cell reconstitution and coordinated B/T cell dysregulation in some PWH. Conversely, CD22 expression, which is uniquely downregulated among high-titer PWH on innate like B cells, showed the opposite pattern, displaying strong negative correlations with these same T cell activation markers **(Figure 6E)**. This divergence points towards a potential mechanism that segregates high-and low-titer PWH. While low-titer PWH were characterized by CD11c and lack of class switching in the presence of T cell activation, high-titer PWH may be able to successfully adapt to this inflammatory environment by downregulating CD22 on non-traditional populations to effectively respond to antigen challenge.

At ≥90 days post-exposure (**Figure 6F**), CD11c-associated B cell features remained positively correlated with T cell activation markers, including CD4 HLA-DR **(Figure 6G)** and CD8 CD25, GLUT1 **(Figure 6H)**, and KLRG1. These findings suggest that persistent immune activation remains associated with B cell remodeling at later post-exposure intervals. However, CD11c-associated B cell features were no longer strongly correlated with total CD4 T cell frequencies **(Figure 6I)**, indicating that relationships between T cell abundance and B cell activation may vary with time since antigen exposure. CD43 expression, which was associated with low Ab titers in PWoH, was negatively correlated with HLA-DR expression on T cells **(Figure 6J)**. These findings suggest that Ab maintenance may be impaired through distinct immune states in PWH and PWoH. In PWH, low titers were associated with a hyper-inflammatory profile marked by coordinated B and T cell activation, whereas in PWoH, low titers were associated with comparatively lower T cell activation and CD43⁺ IgD⁺/MZ-like B cells, suggesting retention of a less differentiated or innate-like B cell state.

## Discussion

Understanding how immune-dysregulated populations develop and maintain humoral immunity after antigen exposure is critical for improving vaccine strategies. Here, we show that low SARS-CoV-2 Ab titers in PWH were associated with broad remodeling of the B cell compartment. Low-titer PWH sampled within 90 days post-exposure displayed expansion of CD11c⁺ and IgM-skewed B cell populations with reduced expression of regulatory markers, whereas those sampled ≥90 days post-exposure showed more heterogeneous alterations involving atypical and memory B cell compartments. These B cell features correlated with T cell activation and differentiation phenotypes, suggesting that impaired Ab induction and maintenance in PWH reflects coordinated immune dysregulation rather than isolated defects in antigen-specific B cell frequency or conventional HIV clinical metrics.

In the present study, we show that CD11c+ atypical B cell expansion is one of the most important features associated with poor Ab titer induction among PWH early post-challenge. These cells, expanded in PWH and other immune-dysregulated cohorts including autoimmunity, aging, and chronic infections^71,83–88^, paradoxically comprise a large portion of spike-specific B cells following SARS-CoV-2 infection and vaccination in HIV-individuals^46,89–91^, though as a transient population not contributing appreciably to long-term memory^89,91^. In contrast, we previously found that solid organ transplant recipients who seroconverted after three vaccine doses had elevated CD11c+ B cells, though with significantly lower Ab titers than PWH^6,92^. We therefore hypothesize that CD11c+ B cells may represent an alternative salvage pathway for mounting Ab responses under profound immunosuppression or dysregulation; while these cells efficiently provide short bursts of proliferative, antigen-specific activity, our data and others suggest they are less productive at generating the high, durable Ab titers maintained long-term in healthy individuals.

A critical determinant of long-term protection against reinfection and severe disease is the maintenance of protective Ab titers, which relies primarily on long-lived plasma cells (LLPCs) that reside in the bone marrow and sustain Ab production long after initial antigen exposure^93–96^. In the present study, PWH who failed to maintain protective Ab titers exhibited heterogeneous peripheral B cell phenotypes, most notably expanded populations of double negative, class-switched memory B cells. This finding aligns with recent reports demonstrating that memory B cells in PWH responding to secondary SARS-CoV-2 exposure preferentially differentiate through extrafollicular pathways rather than canonical germinal center reactions^7,10^. Our findings from early low-titer PWH also supports this interpretation, with CD11c expression, a characteristic marker of extrafollicular B cell activation, leading to decreased GC activity necessary for LLPC development. Collectively, these observations suggest a mechanistic model whereby skewing toward extrafollicular differentiation limits LLPC generation in certain PWH, thereby compromising both the magnitude and durability of Ab responses. Future work should delineate the drivers of CD11c+ atypical B cell expansion in immune dysregulation and explore whether these cells can be therapeutically targeted to enhance vaccine responses in vulnerable populations.

Despite CD4 recovery on ART, it has been well established that many PWH retain altered T cell profiles, including reduced circulating Tfh populations, persistent activation, and exhaustion-associated phenotypes^10,31,80,97–99^. Incomplete restoration of the CD4 compartment has been associated with broad B cell perturbations, including higher frequencies of plasmablasts and activated B cells, a reduced naïve compartment necessary for effective humoral responses, and lower B cell receptor diversity hypothesized to be due to decreased or dysfunctional cTfhs^39,100–102^. Indeed, we show that PWH with lower Ab titers have, on average, lower CD4 counts immediately post-antigen exposure and broad T cell activation that is highly correlated with IgM+, CD11c+ B cell phenotypes. These findings suggest that ongoing T cell dysregulation despite CD4 count recovery favors noncanonical or extrafollicular B cell differentiation that is not best suited for inducing effective, long-term humoral immunity

Several limitations should be considered. The cohort size was modest, particularly after stratification by Ab titer group and post-exposure interval, and participants had heterogeneous vaccine and infection histories. PWH and PWoH were not balanced by sex, limiting our ability to define sex-specific immune signatures. Although sensitivity analyses adjusting for age, sex, BMI, and days since symptom onset showed that most significant B cell subset differences remained significant, the cohort was not powered to comprehensively evaluate interactions between biological sex, HIV status, and Ab titer group. Our analyses were performed in peripheral blood and therefore cannot directly evaluate germinal center reactions or LLPCs. In addition, high-dimensional analyses and B/T cell correlations were exploratory and should be interpreted as hypothesis-generating rather than predictive or causal. Despite these limitations, our findings highlight that low Ab titers in PWH are associated with coordinated B and T cell dysregulation that is not captured by CD4 count or viral suppression alone.

Overall, this study suggests that the durability of SARS-CoV-2 humoral immunity in PWH is shaped by the broader immune landscape established during chronic HIV infection. Identifying the inflammatory and cellular pathways that sustain atypical B cell differentiation despite ART-mediated immune reconstitution may inform vaccine strategies designed to promote durable Ab responses in PWH and other immune-dysregulated populations. Future mechanistic studies will be needed to determine whether modulating these pathways can improve the generation of long-lived memory and plasma cell compartments after vaccination.

## Materials and Methods

### Cohort Description

Participants were enrolled in four arms: COVID+HIV+, COVID+HIV-, COVID-HIV+ and COVID-HIV-^41^. Participants who identified as COVID+ must have been infected with SARS-CoV-2 within 4 weeks of enrolling. COVID-participants must have never tested positive for COVID and believed they never had COVID. Participants in the COVID+HIV+, COVID+HIV-arms completed blood draws at both 1 and 4 months post infection and participants in the COVID-HIV+ and COVID-HIV-arms completed one blood draw at either 0 or 3 months post enrolling. All participants were over 18 years old.

Any individuals in the COVID-HIV+ and COVID-HIV-arms who had positive anti-N IgG levels (indicating prior COVID infection) from the study assays were excluded from the study. Additionally, participants who were reinfected with COVID between the month 1 and month 4 timepoints were excluded from the month 4 population.

### Sex as a biological variable

Our study examined both males and females, but the cohort was not powered to comprehensively evaluate sex-specific immune signatures. However, sensitivity analyses showed that most significant B cell subset differences remained significant after adjusting for sex, BMI, age, and days since symptom onset.

### Serum Antibody Quantification

Serum for antibody quantification was drawn by a mobile phlebotomist into a serum separator tube (SST), centrifuged in the field, and shipped overnight to a central biorepository at Rush University. Occasionally, up to 2-3 days elapsed between phlebotomy and processing due to shipping company delays. Serum was frozen at -80°C until quantification. Previously frozen serum samples were thawed and antibodies quantified with a validated assay using commercial plates manufactured by Meso Scale Discovery (Rockville, Maryland) including the following IgG panels: V-PLEX COVID-19 Coronavirus Panel 2 IgG Kit (9-plex, Catalog No. K15369U-2) - 229E, HKU1, NL63, OC43, CoV-1 Spike, WT N, NTD, RBD, Spike. SARS-CoV-2 Panel 25 IgG - WT, Alpha, Beta, Delta, BA.1, BA.2, BA.3, IHU Spike. SARS-CoV-2 Panel 27 IgG - WT, Beta, Delta, BA.2, BA.3, BA.4, BA.5 Spike. Plates were run on an MESO QuickPlex SQ. Results were analyzed using MSD Discovery Workbench 4.0.12 and titers were converted to WHO-standardized binding antibody units (BAU)/mL. Limit of detection and cutoff was calculated using pre-2020 samples. Month 1 and 4 samples from the same participant were always ran on the same sample plate for all antibodies. All the results were calculated by back-fitting to a same standard curve.

### Ex Vivo Spectral Flow Cytometry and Probe Preparation

PBMCs were analyzed by spectral flow cytometry using previously defined methods^9^. In brief, antigen-specific B cell probes were generated from biotinylated full-length spike protein mixed at a 4:1 protein-to-fluorochrome molar ratio with streptavidin-FITC and streptavidin-BV650 (for spike), calculating ratios based strictly on protein monomer and streptavidin molecular weights. PBMCs stored in liquid nitrogen were thawed using the CryoThaw (“Thawsome”; Medax International, Inc., Salt Lake City, UT) and washed twice with RPMI. Cells were then divided and plated separately for B cell and T cell panel staining, washed in PBS, and stained with BioLegend Live/Dead Aqua Fixable Viability Dye and BD Fc Block for 10 min at RT. Surface staining was performed in 20% BD Horizon Brilliant Stain Buffer in PBS with the antibody cocktail and probes for 20 min at RT. Cells were fixed and permeabilized using the eBioscience FoxP3/Transcription Factor Staining Kit for 20 min at RT, followed by intracellular staining in 1× Permeabilization/Wash buffer for 20 min at RT. Cells were resuspended in 1% paraformaldehyde and acquired on a 4-laser Cytek Aurora spectral flow cytometer. Detailed antibody panel configurations including clones, fluorochromes, manufacturers, and dilutions are provided in **Tables S4 and S5**.

### Statistics

All statistical analyses were performed in GraphPad Prism (version 10.5.0) or R (version 2025.09.1+401). Data are presented as mean ± SEM unless otherwise noted in figure legends. A p-value less than 0.05 was considered statistically significant. Detailed experimental workflows, assay conditions, antibody panels, and custom computational workflows are provided in the Supplemental Methods.

Additional materials and methods can be found in the supplemental materials.

### Study Approval

This study was approved by the Johns Hopkins institutional review board (IRB00278774) and all participants provided written informed consent.

## Data Availability

All data needed to evaluate the conclusions in this paper are present in the manuscript, the Supplemental Materials, and all data and analytic code are available upon request.

## Author Contributions

KRR, AARA, and EAT conceptualized the study. KRR, SBW, and DES developed the methodology. KRR, SBW, AJ, NGM, and DES contributed to investigation and data curation. KRR and DES performed formal analysis. KRR and EAT prepared the visualizations. MJP, ALL, AARA, and EAT acquired funding; MJP, ALL, and AARA provided resources. AJ and NGM contributed to project administration. MJP, ALL, ALC, AARA, and EAT supervised the study. KRR and EAT wrote the original draft of the manuscript. All authors contributed to review and editing of the manuscript and approved the final version.

## Acknowledgments

We would like to acknowledge and thank all study participants for their time and samples. The authors would like to thank all the community members, providers, staff, and other individuals who helped disseminate information about the study, including the Long COVID Justice Network; AIDS Action Baltimore; SisterLove; POZ magazine; Somos Baltimore Latino; the COVID Advocates Advisory Board; The Cranky Queer Newsletter; the Johns Hopkins Institute for Clinical and Translational Research Hopkins Opportunity for Participants Engagement registry; the DC HIV Cohort; and the numerous physicians, researchers, and other individuals who referred participants to the study. We would like to acknowledge the blood processing teams at Rush University and Jia Fu for performing MSD analysis.

## Funding

American Foundation for AIDS Research 110180-69-RSCV.

Johns Hopkins University Center for AIDS Research P30AI094189 (AARA, EAT)

National Institute of Allergy and Infectious Diseases K08AI143391 (AARA)

National Institute of Allergy and Infectious Diseases K22AI175398 (EAT)

## References

1. COVID-19 vaccines | WHO COVID-19 dashboard. Accessed July 6, 2026. https://data.who.int/dashboards/covid19/vaccines

2. COVID-19 cases | WHO COVID-19 dashboard. Accessed July 6, 2026. https://data.who.int/dashboards/covid19/cases

3. Lapuente D, Winkler TH, Tenbusch M. B-cell and antibody responses to SARS-CoV-2: infection, vaccination, and hybrid immunity. Cell Mol Immunol. 2024;21(2):144–158. doi:10.1038/s41423-023-01095-w

4. Bozhkova M, Raycheva R, Petrov S, Dudova D, Kalfova T, Murdjeva M, Taskov H, Shivarov V. Humoral and Memory B Cell Responses Following SARS-CoV-2 Infection and mRNA Vaccination. Vaccines. 2025;13(8):799. doi:10.3390/vaccines13080799

5. Dan JM, Mateus J, Kato Y, Hastie KM, Yu ED, Faliti CE, Grifoni A, Ramirez SI, Haupt S, Frazier A, Nakao C, Rayaprolu V, Rawlings SA, Peters B, Krammer F, Simon V, Saphire EO, Smith DM, Weiskopf D, Sette A, Crotty S. Immunological memory to SARS-CoV-2 assessed for up to 8 months after infection. Science. 2021;371(6529):eabf4063. doi:10.1126/science.abf4063

6. Thompson EA, Figueroa A, Roznik K, Skinner NE, Dhakal S, Li S, Biavati L, Sena LA, Stoddart L, Redinger K, Warner SB, Klein SL, Rouphael N, Blankson JN, Eby Y, Leone RD, Heeger PS, Robien MA, Larsen CP, Pearce EL, Pearce EJ, Ji H, Karaba AH, Segev DL, Tobian AAR, Werbel WA, Cox AL, Bailey JR. Balancing lipid synthesis and oxidation promotes B cell response to vaccination during immunosuppression. J Clin Investig. Published online 2026. doi:10.1172/jci205170

7. Polvere J, Fabbiani M, Pastore G, Rancan I, Rossetti B, Durante M, Zirpoli S, Morelli E, Pettini E, Lucchesi S, Fiorino F, Tumbarello M, Ciabattini A, Montagnani F, Medaglini D. B cell response after SARS-CoV-2 mRNA vaccination in people living with HIV. Commun Med. 2023;3(1):13. doi:10.1038/s43856-023-00245-5

8. Faliti CE, Van TTP, Anam FA, Cheedarla N, Williams ME, Mishra AK, Usman SY, Woodruff MC, Kraker G, Runnstrom MC, Kyu S, Sanz D, Ahmed H, Ghimire M, Morrison-Porter A, Quehl H, Haddad NS, Chen W, Cheedarla S, Neish AS, Roback JD, Antia R, Hom J, Tipton CM, Lindner JM, Ghosn E, Khurana S, Scharer CD, Khosroshahi A, Lee FEH, Sanz I. Disease-associated B cells and immune endotypes shape adaptive immune responses to SARS-CoV-2 mRNA vaccination in human SLE. Nat Immunol. 2025;26(1):131–145. doi:10.1038/s41590-024-02010-9

9. Bergman P, Wullimann D, Gao Y, Borgström EW, Norlin AC, Enoksson SL, Aleman S, Ljunggren HG, Buggert M, Smith CIE. Elevated CD21low B Cell Frequency Is a Marker of Poor Immunity to Pfizer-BioNTech BNT162b2 mRNA Vaccine Against SARS-CoV-2 in Patients with Common Variable Immunodeficiency. J Clin Immunol. 2022;42(4):716–727. doi:10.1007/s10875-022-01244-2

10. Krause R, Snyman J, Shi-Hsia H, Muema D, Karim F, Ganga Y, Ngoepe A, Zungu Y, Gazy I, Bernstein M, Khan K, Mazibuko M, Mthabela N, Ramjit D, Archary M, Dullabh KJ, Giandhari J, Goulder P, Harling G, Harrichandparsad R, Herbst K, Jeena P, Khoza T, Klein N, Madansein R, Marakalala M, Moshabela M, Naidoo K, Ndhlovu Z, Nyamande K, Padayatchi N, Patel V, Smit T, Steyn A, Limbo O, Jardine J, Sok D, Wilson IA, Hanekom W, Sigal A, Kløverpris H, Ndung’u T, Leslie A. HIV skews the SARS-CoV-2 B cell response towards an extrafollicular maturation pathway. eLife. 2022;11:e79924. doi:10.7554/elife.79924

11. Moyon Q, Sterlin D, Miyara M, Anna F, Mathian A, Lhote R, Ghillani-Dalbin P, Breillat P, Mudumba S, Alba S de, Cohen-aubart F, Haroche J, Pha M, Boutin THD, Chaieb H, Flores PM, Charneau P, Gorochov G, Amoura Z. BNT162b2 vaccine-induced humoral and cellular responses against SARS-CoV-2 variants in systemic lupus erythematosus. Ann Rheum Dis. 2022;81(4):575–583. doi:10.1136/annrheumdis-2021-221097

12. Schulz E, Hodl I, Forstner P, Hatzl S, Sareban N, Moritz M, Fessler J, Dreo B, Uhl B, Url C, Grisold AJ, Khalil M, Kleinhappl B, Enzinger C, Stradner MH, Greinix HT, Schlenke P, Steinmetz I. CD19+IgD+CD27-Naïve B Cells as Predictors of Humoral Response to COVID 19 mRNA Vaccination in Immunocompromised Patients. Front Immunol. 2021;12:803742. doi:10.3389/fimmu.2021.803742

13. Autran B, Carcelaint G, Li TS, Gorochov G, Blanc C, Renaud M, Durali M, Mathez D, Calvez V, Leibowitch J, Katlama C, Debré P. Restoration of the immune system with anti-retroviral therapy. Immunol Lett. 1999;66(1-3):207–211. doi:10.1016/s0165-2478(98)00159-x

14. Tseng A, Seet J, Phillips EJ. The evolution of three decades of antiretroviral therapy: challenges, triumphs and the promise of the future. Br J Clin Pharmacol. 2015;79(2):182–194. doi:10.1111/bcp.12403

15. Moysi E, Pallikkuth S, Armas LRD, Gonzalez LE, Ambrozak D, George V, Huddleston D, Pahwa R, Koup RA, Petrovas C, Pahwa S. Altered immune cell follicular dynamics in HIV infection following influenza vaccination. J Clin Investig. 2018;128(7):3171–3185. doi:10.1172/jci99884

16. Höft MA, Burgers WA, Riou C. The immune response to SARS-CoV-2 in people with HIV. Cell Mol Immunol. 2024;21(2):184–196. doi:10.1038/s41423-023-01087-w

17. Zicari S, Sessa L, Cotugno N, Ruggiero A, Morrocchi E, Concato C, Rocca S, Zangari P, Manno EC, Palma P. Immune Activation, Inflammation, and Non-AIDS Co-Morbidities in HIV-Infected Patients under Long-Term ART. Viruses. 2019;11(3):200. doi:10.3390/v11030200

18. Chaer FE, Sahly HME. Vaccination in the Adult Patient Infected with HIV: A Review of Vaccine Efficacy and Immunogenicity. Am J Med. 2019;132(4):437–446. doi:10.1016/j.amjmed.2018.12.011

19. Günthard HF, Wong JK, Spina CA, Ignacio C, Kwok S, Christopherson C, Hwang J, Haubrich R, Havlir D, Richman DD. Effect of Influenza Vaccination on Viral Replication and Immune Response in Persons Infected with Human Immunodeficiency Virus Receiving Potent Antiretroviral Therapy. J Infect Dis. 2000;181(2):522–531. doi:10.1086/315260

20. Mena G, García-Basteiro A, Bayas J. Hepatitis B and A vaccination in HIV-infected adults: A review. Hum Vaccines Immunother. 2015;11(11):2582–2598. doi:10.1080/21645515.2015.1055424

21. Couch RB. Editorial Response: Influenza, Influenza Virus Vaccine, and Human Immunodeficiency Virus Infection. Clin Infect Dis. 1999;28(3):548–551. doi:10.1086/515171

22. Kernéis S, Launay O, Turbelin C, Batteux F, Hanslik T, Boëlle PY. Long-term Immune Responses to Vaccination in HIV-Infected Patients: A Systematic Review and Meta-Analysis. Clin Infect Dis. 2014;58(8):1130–1139. doi:10.1093/cid/cit937

23. Tesoriero JM, Swain CAE, Pierce JL, Zamboni L, Wu M, Holtgrave DR, Gonzalez CJ, Udo T, Morne JE, Hart-Malloy R, Rajulu DT, Leung SYJ, Rosenberg ES. COVID-19 Outcomes Among Persons Living With or Without Diagnosed HIV Infection in New York State. JAMA Netw Open. 2021;4(2):e2037069. doi:10.1001/jamanetworkopen.2020.37069

24. Favara G, Barchitta M, Maugeri A, Faro G, Agodi A. HIV infection does not affect the risk of death of COVID-19 patients: A systematic review and meta-analysis of epidemiological studies. J Glob Heal. 2022;12:05036. doi:10.7189/jogh.12.05036

25. Bertagnolio S, Thwin SS, Silva R, Nagarajan S, Jassat W, Fowler R, Haniffa R, Reveiz L, Ford N, Doherty M, Diaz J. Clinical features of, and risk factors for, severe or fatal COVID-19 among people living with HIV admitted to hospital: analysis of data from the WHO Global Clinical Platform of COVID-19. Lancet HIV. 2022;9(7):e486–e495. doi:10.1016/s2352-3018(22)00097-2

26. Santos C de JS e, Fonseca RR de S, Lima SS, Carvalho TM da S, Mercês LF das, Avelino ME de S, Araújo DO de, Freitas FB, Brasil-Costa I, Oliveira-Filho AB, Vallinoto ACR, Machado LFA. Efficacy of COVID-19 Vaccination in People Living with HIV/AIDS in a Northern Brazil: Cross-Sectional Study. Vaccines. 2025;13(3):283. doi:10.3390/vaccines13030283

27. Ambrosioni J, Blanco JL, Reyes-Urueña JM, Davies MA, Sued O, Marcos MA, Martínez E, Bertagnolio S, Alcamí J, Miro JM, Investigators C 19 in H, Ambrosioni J, Blanco JL, Mora L de la, Garcia-Alcaide F, González-Cordón A, Inciarte A, Laguno M, Leal L, Martínez-Chamorro E, Martínez-Rebollar M, Miró JM, Rojas JF, Torres B, Mallolas J, Albiac L, Agöero DL, Bodro M, Cardozo C, Chumbita M, García N, García-Vidal C, Hernández-Meneses MM, Herrera S, Linares L, Moreno A, Morata L, Martínez-Martínez JA, Puerta P, Rico V, Soriano A, Martínez M, Mosquera M del M, Marcos MA, Vila J, Tuset M, Soy D, Vilella A, Almuedo A, Pinazo MJ, Muñoz J. Overview of SARS-CoV-2 infection in adults living with HIV. Lancet HIV. 2021;8(5):e294–e305. doi:10.1016/s2352-3018(21)00070-9

28. Chun HM, Milligan K, Agyemang E, Ford N, Rangaraj A, Desai S, Wilder-Smith A, Vitoria M, Zulu I. A Systematic Review of COVID-19 Vaccine Antibody Responses in People With HIV. Open Forum Infect Dis. 2022;9(11):ofac579. doi:10.1093/ofid/ofac579

29. Costiniuk CT, Singer J, Lee T, Langlois MA, Arnold C, Galipeau Y, Needham J, Kulic I, Jenabian MA, Burchell AN, Shamji H, Chambers C, Walmsley S, Ostrowski M, Kovacs C, Tan DHS, Harris M, Hull M, Brumme ZL, Lapointe HR, Brockman MA, Margolese S, Mandarino E, Samarani S, Vulesevic B, Lebouché B, Angel JB, Routy JP, Cooper CL, Anis AH, Group CS. COVID-19 vaccine immunogenicity in people with HIV. AIDS. 2023;37(1):F1–F10. doi:10.1097/qad.0000000000003429

30. Vergori A, Lepri AC, Cicalini S, Matusali G, Bordoni V, Lanini S, Meschi S, Iannazzo R, Mazzotta V, Colavita F, Mastrorosa I, Cimini E, Mariotti D, Pascale LD, Marani A, Gallì P, Garbuglia A, Castilletti C, Puro V, Agrati C, Girardi E, Vaia F, Antinori A, Amendola A, Baldini F, Bellagamba R, Bettini A, Bordi L, Camici M, Casetti R, Costantini S, Cristofanelli F, D’Alessio C, D’Aquila V, Angelis AD, Zottis FD, Pascale L de, Francalancia M, Fusto M, Gagliardini R, Gramigna G, Grassi G, Grilli E, Grisetti S, Iafrate D, Lapa D, Lorenzini P, Marani A, Masone E, Marongiu S, Mondi A, Notari S, Ottou S, Paulicelli J, Pellegrino L, Pinnetti C, Plazzi MM, Possi A, Sacchi A, Tartaglia E. Immunogenicity to COVID-19 mRNA vaccine third dose in people living with HIV. Nat Commun. 2022;13(1):4922. doi:10.1038/s41467-022-32263-7

31. Yang X, Zhang J, Liu Z, Chen S, Olatosi B, Poland GA, Weissman S, Li X. COVID-19 breakthrough infections among people living with and without HIV: A statewide cohort analysis. Int J Infect Dis. 2024;139:21–27. doi:10.1016/j.ijid.2023.11.029

32. Pensieroso S, Galli L, Nozza S, Ruffin N, Castagna A, Tambussi G, Hejdeman B, Misciagna D, Riva A, Malnati M, Chiodi F, Scarlatti G. B-cell subset alterations and correlated factors in HIV-1 infection. AIDS. 2013;27(8):1209–1217. doi:10.1097/qad.0b013e32835edc47

33. Jiménez M, Pastor L, Urrea V, Concepción MLR de la, Parker E, Fuente-Soro L, Jairoce C, Mandomando I, Carrillo J, Naniche D, Blanco J. A Longitudinal Analysis Reveals Early Activation and Late Alterations in B Cells During Primary HIV Infection in Mozambican Adults. Front Immunol. 2021;11:614319. doi:10.3389/fimmu.2020.614319

34. Frasca D, Pallikkuth S, Pahwa S. Metabolic phenotype of B cells from young and elderly HIV individuals. Immun Ageing. 2021;18(1):35. doi:10.1186/s12979-021-00245-w

35. Peñaloza N, Rubio L, Sandoval TA, Ballesteros-Ramírez R, Cadena A, Gualtero SM, Valderrama-Beltrán SL, Salazar-Vargas J, Mejía F, Cuéllar A, Quijano S. Incomplete B-cell reconstitution in ART-treated people living with HIV is associated with EBV-linked lymphoma progression. Front Immunol. 2026;17:1791480. doi:10.3389/fimmu.2026.1791480

36. Knox JJ, Buggert M, Kardava L, Seaton KE, Eller MA, Canaday DH, Robb ML, Ostrowski MA, Deeks SG, Slifka MK, Tomaras GD, Moir S, Moody MA, Betts MR. T-bet+ B cells are induced by human viral infections and dominate the HIV gp140 response. JCI Insight. 2017;2(8):e92943. doi:10.1172/jci.insight.92943

37. Martínez LE, Comin-Anduix B, Güemes-Aragon M, Ibarrondo J, Detels R, Mimiaga MJ, Epeldegui M. Characterization of unique B-cell populations in the circulation of people living with HIV prior to non-Hodgkin lymphoma diagnosis. Front Immunol. 2024;15:1441994. doi:10.3389/fimmu.2024.1441994

38. Cascino K, Liechti T, Seaberg EC, Stevens KE, Wolinsky SM, Witt MD, Mailliard RB, Roederer M, Bailey J, Thio CL, Cox AL. HIV causes global B-cell dysregulation and restricts HBV-specific B-cell development in an incident HBV cohort. J Clin Investig. 2026;136(11):e203138. doi:10.1172/jci203138

39. Wang L, Vulesevic B, Vigano M, As’sadiq A, Kang K, Fernandez C, Samarani S, Anis AH, Ahmad A, Costiniuk CT. The Impact of HIV on B Cell Compartment and Its Implications for COVID-19 Vaccinations in People with HIV. Vaccines. 2024;12(12):1372. doi:10.3390/vaccines12121372

40. Touizer E, Alrubbayi A, Ford R, Hussain N, Gerber PP, Shum HL, Rees-Spear C, Muir L, Gea-Mallorquí E, Kopycinski J, Jankovic D, Pinder C, Fox TA, Williams I, Mullender C, Maan I, Waters L, Johnson M, Madge S, Youle M, Barber T, Burns F, Kinloch S, Rowland-Jones S, Gilson R, Matheson NJ, Morris E, Peppa D, McCoy LE. Attenuated humoral responses in HIV infection after SARS-CoV-2 vaccination are linked to global B cell defects and cellular immune profiles. bioRxiv. Published online 2022:2022.11.11.516111. doi:10.1101/2022.11.11.516111

41. Márquez NG, Jamal A, Johnston R, Richter EI, Gorbach PM, Vannorsdall TD, Rubin LH, Jennings C, Landay AL, Peluso MJ, Antar AAR. Characterizing Symptoms and Identifying Biomarkers of Long COVID in People With and Without HIV: Protocol for a Remotely Conducted Prospective Observational Cohort Study. JMIR Res Protoc. 2023;12:e47079. doi:10.2196/47079

42. Srivastava K, Carreño JM, Gleason C, Monahan B, Singh G, Abbad A, Tcheou J, Raskin A, Kleiner G, Bakel H van, Sordillo EM, Group PS, Alshammary H, Amoako AA, Andre D, Awawda M, Bermúdez-González MC, Beach KF, Bielak D, Cai GY, Chernet RL, Cognigni C, Chen Y, Eaker LQ, Ferreri ED, Floda DL, Fried M, Hamburger JZ, Jurczyszak D, Kang HM, Lyttle N, Matthews JC, Mauldin J, Mendez WA, Mischka J, Morris S, Mulder LCF, Nabeel I, Nardulli JR, Polanco J, Oostenink A, Rooker A, Russo KT, Salimbangon AB, Saksena MS, Shin AA, Sominsky LA, Stadlbauer D, Sullivan L, Kesteren M van, Yellin T, Wajnberg A, Krammer F, Simon V. SARS-CoV-2-infection-and vaccine-induced antibody responses are long lasting with an initial waning phase followed by a stabilization phase. Immunity. 2024;57(3):587–599.e4. doi:10.1016/j.immuni.2024.01.017

43. Pradenas E, Urrea V, Marfil S, Pidkova T, Aguilar-Gurrieri C, Abancó F, Mateu L, Chamorro A, Grau E, Trigueros M, Carrillo J, Massanella M, Trinité B, Clotet B, Blanco J. Recurrent waning of anti-SARS-CoV-2 neutralizing antibodies despite multiple antigen encounters. J Transl Med. 2025;23(1):783. doi:10.1186/s12967-025-06837-0

44. Andrews N, Tessier E, Stowe J, Gower C, Kirsebom F, Simmons R, Gallagher E, Thelwall S, Groves N, Dabrera G, Myers R, Campbell CNJ, Amirthalingam G, Edmunds M, Zambon M, Brown K, Hopkins S, Chand M, Ladhani SN, Ramsay M, Bernal JL. Duration of Protection against Mild and Severe Disease by Covid-19 Vaccines. N Engl J Med. 2022;386(4):340–350. doi:10.1056/nejmoa2115481

45. Byrne J, Gaillard CM, Gu L, Garcia-Leon A, Casey O, Kenny G, Saini G, Alalwan D, O’Gorman T, O’Regan S, Doran P, Feeney ER, O’Halloran JA, Horgan M, Cotter AG, Barra E de, Sadlier C, Landay A, Gautier V, Mallon PWG. Specific thresholds of circulating antibody titers predict against infection and reduced disease severity in SARS-CoV-2 close contacts. J Immunol. 2025;214(9):2238–2243. doi:10.1093/jimmun/vkaf101

46. Steuten J, Bos AV, Kuijper LH, Claireaux M, Olijhoek W, Elias G, Duurland MC, Jorritsma T, Marsman C, Paul AGA, Vallejo JJG, Gils MJ van, Wieske L, Kuijpers TW, Eftimov F, Ham SM van, Brinke A ten, Consortium T. Distinct dynamics of antigen-specific induction and differentiation of different CD11c+Tbet+ B-cell subsets. J Allergy Clin Immunol. 2023;152(3):689–699.e6. doi:10.1016/j.jaci.2023.02.020

47. Tangye SG. Do multiple subsets of CD11c+ B cells exist? You (T)-Bet! J Allergy Clin Immunol. 2023;152(3):607–609. doi:10.1016/j.jaci.2023.07.004

48. Sanz I, Wei C, Jenks SA, Cashman KS, Tipton C, Woodruff MC, Hom J, Lee FEH. Challenges and Opportunities for Consistent Classification of Human B Cell and Plasma Cell Populations. Front Immunol. 2019;10:2458. doi:10.3389/fimmu.2019.02458

49. Cascino K, Roederer M, Liechti T. OMIP-068: High-Dimensional Characterization of Global and Antigen-Specific B Cells in Chronic Infection. Cytom Part A. 2020;97(10):1037–1043. doi:10.1002/cyto.a.24204

50. Ma S, Wang C, Mao X, Hao Y. B Cell Dysfunction Associated With Aging and Autoimmune Diseases. Front Immunol. 2019;10:318. doi:10.3389/fimmu.2019.00318

51. Nipper AJ, Smithey MJ, Shah RC, Canaday DH, Landay AL. Diminished antibody response to influenza vaccination is characterized by expansion of an age-associated B-cell population with low PAX5. Clin Immunol. 2018;193:80–87. doi:10.1016/j.clim.2018.02.003

52. Yam-Puc JC, Hosseini Z, Horner EC, Gerber PP, Beristain-Covarrubias N, Hughes R, Lulla A, Rust M, Boston R, Ali M, Fischer K, Simmons-Rosello E, O’Reilly M, Robson H, Booth LH, Kahanawita L, Correa-Noguera A, Favara D, Ceron-Gutierrez L, Keller B, Craxton A, Anderson GSF, Sun XM, Elmer A, Saunders C, Bermperi A, Jose S, Kingston N, Mulroney TE, Piñon LPG, Collaboration CNCB, Chapman MA, Grigoriadou S, MacFarlane M, Willis AE, Patil KR, Spencer S, Staples E, Warnatz K, Buckland MS, Hollfelder F, Hyvönen M, Döffinger R, Parkinson C, Lear S, Matheson NJ, Thaventhiran JED. Age-associated B cells predict impaired humoral immunity after COVID-19 vaccination in patients receiving immune checkpoint blockade. Nat Commun. 2023;14(1):3292. doi:10.1038/s41467-023-38810-0

53. Veri M, Gorlatov S, Li H, Burke S, Johnson S, Stavenhagen J, Stein KE, Bonvini E, Koenig S. Monoclonal antibodies capable of discriminating the human inhibitory Fcγ-receptor IIB (CD32B) from the activating Fcγ-receptor IIA (CD32A): biochemical, biological and functional characterization. Immunology. 2007;121(3):392–404. doi:10.1111/j.1365-2567.2007.02588.x

54. Rankin CT, Veri MC, Gorlatov S, Tuaillon N, Burke S, Huang L, Inzunza HD, Li H, Thomas S, Johnson S, Stavenhagen J, Koenig S, Bonvini E. CD32B, the human inhibitory Fc-γ receptor IIB, as a target for monoclonal antibody therapy of B-cell lymphoma. Blood. 2006;108(7):2384–2391. doi:10.1182/blood-2006-05-020602

55. Takai T. Roles of Fc receptors in autoimmunity. Nat Rev Immunol. 2002;2(8):580–592. doi:10.1038/nri856

56. Trend S, Leffler J, Teige I, Frendéus B, Kermode AG, French MA, Hart PH. FcγRIIb Expression Is Decreased on Naive and Marginal Zone-Like B Cells From Females With Multiple Sclerosis. Front Immunol. 2021;11:614492. doi:10.3389/fimmu.2020.614492

57. Mackay M, Stanevsky A, Wang T, Aranow C, Li M, Koenig S, Ravetch JV, Diamond B. Selective dysregulation of the FcγIIB receptor on memory B cells in SLE. J Exp Med. 2006;203(9):2157–2164. doi:10.1084/jem.20051503

58. Liu Y, Gong Y, Qu C, Zhang Y, You R, Yu N, Lu G, Huang Y, Zhang H, Gao Y, Gao Y, Guo X. CD32b expression is down-regulated on double-negative memory B cells in patients with Hashimoto’s thyroiditis. Mol Cell Endocrinol. 2017;440:1–7. doi:10.1016/j.mce.2016.11.004

59. Isaák A, Gergely P, Szekeres Z, Prechl J, Poór G, Erdei A, Gergely J. Physiological up-regulation of inhibitory receptors FcγRII and CR1 on memory B cells is lacking in SLE patients. Int Immunol. 2008;20(2):185–192. doi:10.1093/intimm/dxm132

60. Thibult ML, Mamessier E, Gertner-Dardenne J, Pastor S, Just-Landi S, Xerri L, Chetaille B, Olive D. PD-1 is a novel regulator of human B-cell activation. Int Immunol. 2013;25(2):129–137. doi:10.1093/intimm/dxs098

61. Martin F, Oliver AM, Kearney JF. Marginal Zone and B1 B Cells Unite in the Early Response against T-Independent Blood-Borne Particulate Antigens. Immunity. 2001;14(5):617–629. doi:10.1016/s1074-7613(01)00129-7

62. Appelgren D, Eriksson P, Ernerudh J, Segelmark M. Marginal-Zone B-Cells Are Main Producers of IgM in Humans, and Are Reduced in Patients With Autoimmune Vasculitis. Front Immunol. 2018;9:2242. doi:10.3389/fimmu.2018.02242

63. Palm AKE, Kleinau S. Marginal zone B cells: From housekeeping function to autoimmunity? J Autoimmun. 2021;119:102627. doi:10.1016/j.jaut.2021.102627

64. Cambier JC, Gauld SB, Merrell KT, Vilen BJ. B-cell anergy: from transgenic models to naturally occurring anergic B cells? Nat Rev Immunol. 2007;7(8):633–643. doi:10.1038/nri2133

65. Merrell KT, Benschop RJ, Gauld SB, Aviszus K, Decote-Ricardo D, Wysocki LJ, Cambier JC. Identification of Anergic B Cells within a Wild-Type Repertoire. Immunity. 2006;25(6):953–962. doi:10.1016/j.immuni.2006.10.017

66. Liubchenko GA, Appleberry HC, Holers VM, Banda NK, Willis VC, Lyubchenko T. Potentially autoreactive naturally occurring transitional T3 B lymphocytes exhibit a unique signaling profile. J Autoimmun. 2012;38(4):293–303. doi:10.1016/j.jaut.2011.12.005

67. Clark EA, Giltiay NV. CD22: A Regulator of Innate and Adaptive B Cell Responses and Autoimmunity. Front Immunol. 2018;9:2235. doi:10.3389/fimmu.2018.02235

68. Tsubata T. Ligand Recognition Determines the Role of Inhibitory B Cell Co-receptors in the Regulation of B Cell Homeostasis and Autoimmunity. Front Immunol. 2018;9:2276. doi:10.3389/fimmu.2018.02276

69. Bernard NJ. Double-negative B cells. Nat Rev Rheumatol. 2018;14(12):684–684. doi:10.1038/s41584-018-0113-6

70. Colonna-Romano G, Bulati M, Aquino A, Pellicanò M, Vitello S, Lio D, Candore G, Caruso C. A double-negative (IgD−CD27−) B cell population is increased in the peripheral blood of elderly people. Mech Ageing Dev. 2009;130(10):681–690. doi:10.1016/j.mad.2009.08.003

71. Jenks SA, Cashman KS, Zumaquero E, Marigorta UM, Patel AV, Wang X, Tomar D, Woodruff MC, Simon Z, Bugrovsky R, Blalock EL, Scharer CD, Tipton CM, Wei C, Lim SS, Petri M, Niewold TB, Anolik JH, Gibson G, Lee FEH, Boss JM, Lund FE, Sanz I. Distinct Effector B Cells Induced by Unregulated Toll-like Receptor 7 Contribute to Pathogenic Responses in Systemic Lupus Erythematosus. Immunity. 2018;49(4):725–739.e6. doi:10.1016/j.immuni.2018.08.015

72. Chung MKY, Gong L, Kwong DL, Lee VH, Lee AW, Guan X, Kam N, Dai W. Functions of double-negative B cells in autoimmune diseases, infections, and cancers. EMBO Mol Med. 2023;15(9):EMMM202217341. doi:10.15252/emmm.202217341

73. Li Y, Li Z, Hu F. Double-negative (DN) B cells: an under-recognized effector memory B cell subset in autoimmunity. Clin Exp Immunol. 2021;205(2):119–127. doi:10.1111/cei.13615

74. Dirks J, Andres O, Paul L, Manukjan G, Schulze H, Morbach H. IgD shapes the pre-immune naïve B cell compartment in humans. Front Immunol. 2023;14:1096019. doi:10.3389/fimmu.2023.1096019

75. Noviski M, Mueller JL, Satterthwaite A, Garrett-Sinha LA, Brombacher F, Zikherman J. IgM and IgD B cell receptors differentially respond to endogenous antigens and control B cell fate. eLife. 2018;7:e35074. doi:10.7554/elife.35074

76. Sabouri Z, Perotti S, Spierings E, Humburg P, Yabas M, Bergmann H, Horikawa K, Roots C, Lambe S, Young C, Andrews TD, Field M, Enders A, Reed JH, Goodnow CC. IgD attenuates the IgM-induced anergy response in transitional and mature B cells. Nat Commun. 2016;7(1):13381. doi:10.1038/ncomms13381

77. Roes J, Rajewsky K. Immunoglobulin D (IgD)-deficient mice reveal an auxiliary receptor function for IgD in antigen-mediated recruitment of B cells. J Exp Med. 1993;177(1):45–55. doi:10.1084/jem.177.1.45

78. Wikén M, Björck P, Axelsson B, Perlmann P. Studies on the Role of CD43 in Human B-Cell Activation and Differentiation. Scand J Immunol. 1989;29(3):353–361. doi:10.1111/j.1365-3083.1989.tb01134.x

79. Wikén M, Björck P, Axelsson B, Perlmann P. Induction of CD43 Expression during Activation and Terminal Differentiation of Human B Cells. Scand J Immunol. 1988;28(4):457–464. doi:10.1111/j.1365-3083.1988.tb01476.x

80. Mu W, Patankar V, Kitchen S, Zhen A. Examining Chronic Inflammation, Immune Metabolism, and T Cell Dysfunction in HIV Infection. Viruses. 2024;16(2):219. doi:10.3390/v16020219

81. Babu H, Ambikan AT, Gabriel EE, Akusjärvi SS, Palaniappan AN, Sundaraj V, Mupanni NR, Sperk M, Cheedarla N, Sridhar R, Tripathy SP, Nowak P, Hanna LE, Neogi U. Systemic Inflammation and the Increased Risk of Inflamm-Aging and Age-Associated Diseases in People Living With HIV on Long Term Suppressive Antiretroviral Therapy. Front Immunol. 2019;10:1965. doi:10.3389/fimmu.2019.01965

82. Hileman CO, Funderburg NT. Inflammation, Immune Activation, and Antiretroviral Therapy in HIV. Curr HIVAIDS Rep. 2017;14(3):93–100. doi:10.1007/s11904-017-0356-x

83. Rubtsov AV, Rubtsova K, Fischer A, Meehan RT, Gillis JZ, Kappler JW, Marrack P. Toll-like receptor 7 (TLR7)–driven accumulation of a novel CD11c+ B-cell population is important for the development of autoimmunity. Blood. 2011;118(5):1305–1315. doi:10.1182/blood-2011-01-331462

84. Karnell JL, Kumar V, Wang J, Wang S, Voynova E, Ettinger R. Role of CD11c+ T-bet+ B cells in human health and disease. Cell Immunol. 2017;321:40–45. doi:10.1016/j.cellimm.2017.05.008

85. Zhang W, Zhang H, Liu S, Xia F, Kang Z, Zhang Y, Liu Y, Xiao H, Chen L, Huang C, Shen N, Xu H, Li F. Excessive CD11c+Tbet+ B cells promote aberrant TFH differentiation and affinity-based germinal center selection in lupus. Proc Natl Acad Sci. 2019;116(37):18550–18560. doi:10.1073/pnas.1901340116

86. Sundling C, Rönnberg C, Yman V, Asghar M, Jahnmatz P, Lakshmikanth T, Chen Y, Mikes J, Forsell MN, Sondén K, Achour A, Brodin P, Persson KEM, Färnert A. B cell profiling in malaria reveals expansion and remodelling of CD11c+ B cell subsets. JCI Insight. 2019;4(9). doi:10.1172/jci.insight.126492

87. Amano E, Sato W, Kimura Y, Kimura A, Lin Y, Okamoto T, Sato N, Yokota T, Yamamura T. CD11chigh B Cell Expansion Is Associated With Severity and Brain Atrophy in Neuromyelitis Optica. Neurol Neuroimmunol Neuroinflammation. 2024;11(2):e200206. doi:10.1212/nxi.0000000000200206

88. Maul RW, Catalina MD, Kumar V, Bachali P, Grammer AC, Wang S, Yang W, Hasni S, Ettinger R, Lipsky PE, Gearhart PJ. Transcriptome and IgH Repertoire Analyses Show That CD11chi B Cells Are a Distinct Population With Similarity to B Cells Arising in Autoimmunity and Infection. Front Immunol. 2021;12:649458. doi:10.3389/fimmu.2021.649458

89. Kardava L, Rachmaninoff N, Lau WW, Buckner CM, Trihemasava K, Blazkova J, Assis FL de, Wang W, Zhang X, Wang Y, Chiang CI, Narpala S, McCormack GE, Liu C, Seamon CA, Sneller MC, O’Connell S, Li Y, McDermott AB, Chun TW, Fauci AS, Tsang JS, Moir S. Early human B cell signatures of the primary antibody response to mRNA vaccination. Proc Natl Acad Sci. 2022;119(28):e2204607119. doi:10.1073/pnas.2204607119

90. Sokal A, Chappert P, Barba-Spaeth G, Roeser A, Fourati S, Azzaoui I, Vandenberghe A, Fernandez I, Meola A, Bouvier-Alias M, Crickx E, Beldi-Ferchiou A, Hue S, Languille L, Michel M, Baloul S, Noizat-Pirenne F, Luka M, Mégret J, Ménager M, Pawlotsky JM, Fillatreau S, Rey FA, Weill JC, Reynaud CA, Mahévas M. Maturation and persistence of the anti-SARS-CoV-2 memory B cell response. Cell. 2021;184(5):1201–1213.e14. doi:10.1016/j.cell.2021.01.050

91. Assis FL de, Hoehn KB, Zhang X, Kardava L, Smith CD, Merhebi OE, Buckner CM, Trihemasava K, Wang W, Seamon CA, Chen V, Schaughency P, Cheung F, Martins AJ, Chiang CI, Li Y, Tsang JS, Chun TW, Kleinstein SH, Moir S. Tracking B cell responses to the SARS-CoV-2 mRNA-1273 vaccine. Cell Rep. 2023;42(7):112780. doi:10.1016/j.celrep.2023.112780

92. Haidar G, Agha M, Bilderback A, Lukanski A, Linstrum K, Troyan R, Rothenberger S, McMahon DK, Crandall MD, Sobolewksi MD, Enick PN, Jacobs JL, Collins K, Klamar-Blain C, Macatangay BJC, Parikh UM, Heaps A, Coughenour L, Schwartz MB, Dueker JM, Silveira FP, Keebler ME, Humar A, Luketich JD, Morrell MR, Pilewski JM, McDyer JF, Pappu B, Ferris RL, Marks SM, Mahon J, Mulvey K, Hariharan S, Updike GM, Brock L, Edwards R, Beigi RH, Kip PL, Wells A, Minnier T, Angus DC, Mellors JW. Prospective Evaluation of Coronavirus Disease 2019 (COVID-19) Vaccine Responses Across a Broad Spectrum of Immunocompromising Conditions: the COVID-19 Vaccination in the Immunocompromised Study (COVICS). Clin Infect Dis. 2022;75(1):e630–e644. doi:10.1093/cid/ciac103

93. Nutt SL, Hodgkin PD, Tarlinton DM, Corcoran LM. The generation of antibody-secreting plasma cells. Nat Rev Immunol. 2015;15(3):160–171. doi:10.1038/nri3795

94. Slifka MK, Antia R, Whitmire JK, Ahmed R. Humoral Immunity Due to Long-Lived Plasma Cells. Immunity. 1998;8(3):363–372. doi:10.1016/s1074-7613(00)80541-5

95. Amanna IJ, Carlson NE, Slifka MK. Duration of Humoral Immunity to Common Viral and Vaccine Antigens. N Engl J Med. 2007;357(19):1903–1915. doi:10.1056/nejmoa066092

96. Manz RA, Thiel A, Radbruch A. Lifetime of plasma cells in the bone marrow. Nature. 1997;388(6638):133–134. doi:10.1038/40540

97. Okoye AA, Picker LJ. CD4+ T-cell depletion in HIV infection: mechanisms of immunological failure. Immunol Rev. 2013;254(1):54–64. doi:10.1111/imr.12066

98. Pallikkuth S, Parmigiani A, Silva SY, George VK, Fischl M, Pahwa R, Pahwa S. Impaired peripheral blood T-follicular helper cell function in HIV-infected nonresponders to the 2009 H1N1/09 vaccine. Blood. 2012;120(5):985–993. doi:10.1182/blood-2011-12-396648

99. Aksak-Wąs B, Skonieczna-Żydecka K, Parczewski M, Hrynkiewicz R, Lewandowski F, Serwin K, Mielczak K, Majchrzak A, Bruss M, Niedźwiedzka-Rystwej P. Immunorecovered but Exhausted: Persistent PD-1/PD-L1 Expression Despite Virologic Suppression and CD4 Recovery in PLWH. Biomedicines. 2025;13(8):1885. doi:10.3390/biomedicines13081885

100. Moir S, Fauci AS. Insights into B cells and HIV-specific B-cell responses in HIV-infected individuals. Immunol Rev. 2013;254(1):207–224. doi:10.1111/imr.12067

101. Lu X, Zhang X, Cheung AKL, Moog C, Xia H, Li Z, Wang R, Ji Y, Xia W, Liu Z, Yuan L, Wang X, Wu H, Zhang T, Su B. Abnormal Shift in B Memory Cell Profile Is Associated With the Expansion of Circulating T Follicular Helper Cells via ICOS Signaling During Acute HIV-1 Infection. Front Immunol. 2022;13:837921. doi:10.3389/fimmu.2022.837921

102. Wong CS, Buckner CM, Lage SL, Pei L, Assis FL, Dahlstrom EW, Anzick SL, Virtaneva K, Rupert A, Davis JL, Zhou T, Laidlaw E, Manion M, Galindo F, Anderson M, Seamon CA, Sneller MC, Lisco A, Deleage C, Pittaluga S, Moir S, Sereti I. Rapid Emergence of T Follicular Helper and Germinal Center B Cells Following Antiretroviral Therapy in Advanced HIV Disease. Front Immunol. 2021;12:752782. doi:10.3389/fimmu.2021.752782

