## Supplemental Materials for "An altered B cell compartment distinguishes low antibody responders among people with HIV"

Redinger et al

### Materials and Methods

#### Sample Processing

Blood was collected by mobile phlebotomy, processed in the field, and shipped to a Rush University lab for clinical labs, processing, and frozen storage. The time between blood draw and processing varied between 20 hours and up to 3 days.

#### Spectral Flow Cytometry Analysis

FCS files were analyzed using the OMIQ software from Dotmatics (manual gating strategies are detailed in Figs. S2 and S6). B cells were identified by sequential gating on lymphocytes (FSC-A vs SSC-A), singlets (FSC-A vs FSC-H), dump channel negative (Live dead Aqua$^-$,CD3$^-$,CD14$^-$), CD19$^+$ and/or CD20$^+$ followed by data cleaning using flowAI. All files were then normalized using Cycombine to account for any batch effects. B cell frequencies were obtained using the gating strategy defined previously **(Figure S3A)**, and median fluorescent intensities (MFIs) were exported and plotted in GraphPad Prism (version 10.5.0) or used for downstream analyses in R (version 2025.09.1+401). T cells were identified using a separate flow cytometry panel and identified by sequential gating on lymphocytes (FSC-A vs SSC-A), singlets (FSC-A vs FSC-H), live cells (Live dead Aqua$^-$), and specific subset markers defined in Figure S6A. Frequencies and MFIs were exported for downstream analyses in R (version 2025.09.1+401).

#### Statistical Analysis of B Cell Frequencies

Initial unadjusted analyses were performed in GraphPad Prism (version 10.5.0). For each B cell subset, Kruskal-Wallis tests compared frequencies across the four groups (HCs-High, HCs-Low, PLWH-High, PLWH-Low), followed by Dunn's multiple comparison tests (method = "none") to evaluate all pairwise group differences independently for the early and late time points.

To adjust for potential confounding by demographic or clinical variables, covariate-adjusted analyses were performed in R (version 2025.09.1+401). For each B cell subset, we implemented a rank-based ANCOVA procedure: (1) frequency values were converted to ranks; (2) ranks were regressed on age, sex, and BMI using ordinary least squares regression; (3) residuals from these models represent covariate-adjusted ranks; (4) Dunn's post-hoc tests were applied to the residuals using the **dunnTest** function from the FSA package with method = "none" to obtain adjusted pairwise p-values. This approach maintains the distribution-free properties of non-parametric tests while adjusting for covariates. Forest plots present both unadjusted (Prism) and adjusted (R) p-values for comparisons where at least one p-value was < 0.05.

**High Dimensional Analyses**

All high dimensional analyses were performed in R (version 2025.09.1+401) using the following packages: **MASS**, **caret**, **ggplot2**, **corrplot**, **pysch, rstatix,** and **Hmisc**. All analyses were performed separately for early and late timepoints. A random seed (123) was set to ensure reproducibility. R code for data analysis was developed with assistance from Claude Sonnet 4.5 (Anthropic, 2025). All generated code was independently verified and tested by the authors.

**Linear Discriminant Analysis and Variable Selection**

Linear discriminant analysis was employed using the lda function from the **MASS** package (v7.3-60.0.1) to identify the most discriminatory B cell phenotypic features between response groups. Prior to LDA, data preprocessing was performed using the **caret** package (v7.0.1). Continuous variables were mean-centered and scaled using z-score normalization, while categorical variables (vaccine number and sex) were converted to dummy variables. Near-zero variance predictors were identified and removed using the nearZeroVar function from caret. A detailed list of variables used for analysis are available in **Table S2**.

Variable importance was calculated using Cohen's d effect size, which quantified the discriminatory power of each variable for separating each response group from all others. For each variable, the maximum Cohen's d across all response groups was used as the overall importance score. Variables with importance scores exceeding a threshold of 1.0 were retained for subsequent analyses. LDA results were visualized using multiple approaches implemented with **ggplot2** (v3.5.2). Scatter plots of the first two linear discriminants (LD1 and LD2) were created with 95% confidence ellipses for each response group using stat_ellipse, and heatmaps of mean variable expression across groups were generated with hierarchical clustering applied to both variables (rows) and groups (columns).

**Hierarchical Clustering and Composite Score Generation**

To reduce redundancy among high-importance variables and create interpretable composite measures, we performed hierarchical clustering of correlated variables. Spearman correlation matrices were calculated for all retained high-importance variables using the base R cor function. Correlation matrices were visualized using the **corrplot** package (v0.95).

Hierarchical clustering was performed using complete linkage on a distance matrix derived from correlation coefficients, where distance = 1 - |correlation|. Dendrograms were generated to visualize variable relationships, and clusters were defined by cutting the dendrogram at a height of 0.7. Variable cluster assignments were saved for interpretation and downstream analyses.

For each identified cluster, composite scores were generated using standardized z-scores (mean = 0, standard deviation = 1) using the scale() function to ensure equal weighting regardless of original measurement scales. For clusters containing multiple variables (≥2 variables with $|r| \geq 0.3$), composite scores were calculated as the arithmetic mean of the z-scored variables within that cluster using the rowMeans() function, with na.rm = TRUE to handle missing values. For singleton clusters (single variables not meeting the correlation threshold with any other variable), the standardized z-score of that individual variable was used directly as the composite score, maintaining consistency in the scale across all composites. This approach reduced the dimensionality of the dataset while preserving biological information. All composite scores were calculated separately for early and late timepoint variables. Composite score distributions across response groups were visualized using boxplots with overlaid individual data points, created with **ggplot2** (v3.5.2).

**Statistical Comparison of Composite Scores**

To assess whether composite scores significantly differed between antibody response groups, we employed non-parametric statistical testing using the **rstatix** package (v0.7.2. For each composite score, the Kruskal-Wallis test was used as an omnibus test to evaluate overall differences among the multiple response groups. Only composite scores showing significant Kruskal-Wallis results ($p < 0.05$) were subjected to post-hoc pairwise comparisons.

For significant composite scores, pairwise comparisons were then conducted using Dunn's test with false discovery rate (FDR) correction for multiple comparisons. Pairwise comparisons with FDR-adjusted $p < 0.05$ were considered statistically significant. Composite scores that did not show significant overall group differences in the Kruskal-Wallis test were not subjected to pairwise testing. This conditional testing approach reduces the multiple testing burden while maintaining appropriate control of family-wise error rates. Composite score distributions across response groups were visualized using boxplots with overlaid individual data points, created with ggplot2 (v3.5.2).

**Correlation Analysis Between B Cell and T Cell Variables**

To investigate relationships between B cell phenotypic composite scores and T cell functional measurements, we performed correlation analyses between significant B cell composite scores (identified from the statistical testing above) and normalized T cell flow cytometry variables (full list of variables available in **Table S3)**. T cell data were matched to B cell samples using unique file identifiers and were preprocessed identically to B cell data: continuous variables were centered and scaled, near-zero variance features were removed, and only complete cases were retained for analysis.

Spearman rank correlations were calculated between all significant B cell composite scores and all T cell variables using the **Hmisc** package (v5.2.3). The resulting correlation coefficient matrix and associated p-values were extracted. To account for multiple hypothesis testing, p-values were adjusted using the false discovery rate (FDR) method. Correlations were considered significant at FDR < 0.05.

Significant correlation patterns were visualized using dot plot heatmaps. Prior to plotting, hierarchical clustering (average linkage) was performed independently on both B cell composites and T cell variables based on their correlation profiles to group similar patterns together. In the dot plots, correlation coefficients (ρ) were represented by dot color, while dot size represented statistical significance ($-\log_{10}$(adjusted p-value)).

Tables

**Table S1. Cohort demographics**

| Characteristic | Overall<br>N = 105 | PLWH<br>N = 39 | HCs<br>N = 66 | p-value[1] |
|---|---|---|---|---|
| **Age** | | | | 0.413 |
| 20-39 | 45 (43%) | 14 (36%) | 31 (47%) | |
| 40-59 | 46 (44%) | 18 (46%) | 28 (42%) | |
| 60+ | 14 (13%) | 7 (18%) | 7 (11%) | |
| **Sex** | | | | **<0.001** |
| Male | 59 (56%) | 32 (82%) | 27 (41%) | |
| Female | 46 (44%) | 7 (18%) | 39 (59%) | |
| **BMI** | | | | 0.673 |
| Healthy (18.5-24.9) | 38 (36%) | 12 (31%) | 26 (39%) | |
| Overweight (25.0-29.9) | 37 (35%) | 15 (38%) | 22 (33%) | |
| Obese (≥ 30.0) | 30 (29%) | 12 (31%) | 18 (27%) | |
| **Last Antigen Exposure** | | | | 0.640 |
| Vaccine | 23 (22%) | 10 (26%) | 13 (20%) | |
| Infection | 82 (78%) | 29 (74%) | 53 (80%) | |
| **Number of Vaccines** | | | | 0.922 |
| 1 | 3 (3%) | 1 (3%) | 2 (3%) | |
| 2 | 42 (40%) | 15 (38%) | 27 (41%) | |
| 3 | 56 (53%) | 21 (54%) | 35 (53%) | |
| 4 | 4 (4%) | 2 (5%) | 2 (3%) | |
| 5 | 0 (0%) | 0 (0%) | 0 (0%) | |

| Characteristic | Overall<br>N = 105 | PLWH<br>N = 39 | HCs<br>N = 66 | p-value[1] |
|---|---|---|---|---|
| **CD4 Count[2]** | | | | N/A |
| High (≥500) | 19 (63%) | 19 (63%) | - | |
| Low (<500) | 11 (37%) | 11 (37%) | - | |

[1]Chi-squared test; Fisher's exact test for variables with expected cell counts <5

[2]CD4 counts were available for 30 out of 39 samples for PLWH based on availability of fresh blood

**Table S2. Linear discriminant analysis (LDA) variables for high dimensional B cell profiling**

| B cell subsets | Marker MFIs | Clinical parameters |
|---|---|---|
| Transitional | PD-1 | Age |
| T3 | CD32 | BMI |
| T2-MZP | CD22 | Sex |
| B1 B cells | CD43 | Vaccine Number |
| CD11c+ B cells | CD39 | Last exposure type |
| TBET- CD11c | CD11c | Days since symptom onset |
| Naive | CD10 | |
| Resting Naive | FcRL5 | |
| Activated Naive | TBET | |
| DN | CPT1a | |
| DN1 | Glut1 | |
| DN3 | CXCR5 | |
| Classical Memory | CXCR3 | |
| IgG MBC | IgM | |
| IgM MBC | IgG | |
| IgM/IgG neg SwM | IgD | |
| actMBC | | |
| resMBC | | |
| IgD+ CD27+ | | |
| IgD only MBC | | |
| MZ like | | |
| Spike+ | | |

**Table S3. T cell variables for B cell composite correlations**

| T cell frequencies | Marker MFIs (on bulk CD4 and CD8 T cells) |
|---|---|
| Total CD4 | CPT1a |
| Total CD8 | Glut1 |
| Tregs | PD-1 |
| CD4 Tcm | CD11c |
| CD4 naive | CD25 |
| CD4 Tem | CD38 |
| CD4 Temra | HLA-DR |
| CD8 Tcm | KLRG1 |
| CD8 naive | Ki67 |
| CD8 Tem | CD27 |
| CD8 Temra | |

**Table S4. B cell flow cytometry panel configuration**

| Antibody | Fluorophore | Clone | Manufacturer | Catalog # |
|---|---|---|---|---|
| **Surface** | | | | |
| CD10 | PE Cy5 | HI10a | BD | 555376 |
| CD24 | BUV395 | ML5 | BD | 566221 |
| CD38 | BUV661 | HIT2 | BD | 565070 |
| IgM | AF700 | MHM88 | Biolegend | 314537 |
| CD21 | BUV496 | B-ly4 | BD | 624283 |
| CD86 | BUV737 | 2331(FUN-1) | BD | 612784 |
| CD22 | BUV805 | HIB22 | BD | 742009 |
| CXCR3 | APC | G025H7 | Biolegend | 353707 |
| PD-1 | BUV615 | EH12.1 | BD | 612991 |
| FcRL5 (CD307e) | BB660 | 509F6 | BD-Crada | 624295 |
| BTLA | BB700 | J168-540 | BD | 746166 |
| CXCR5 | PE Cy7 | J252D4 | Biolegend | 356923 |
| CD19 | APC H7 | H1B19 | BD | 560727 |
| FcγRIIb (CD32) | BV750 | 3D3 | BD | 747110 |
| CD71 | PERCP-Cy5.5 | CY1G4 | Biolegend | 334114 |
| CD36 | BV480 | CLB-IVC7 | BD | 746612 |
| CD11c | BB630 | BU15 | BD-Crada | 624294 |
| CD27 | PE CF594 | M-T271 | BD | 562297 |
| CD39 | BV711 | Tu66 | BD | 563680 |
| CD20 | BV786 | TH7 | BD | 743611 |
| CD3 | BV510 | UCHT1 | BD | 563109 |
| CD14 | BV510 | MφP9 | BD | 563079 |
| IgD | BB790 | IA6-2 | BD-Crada | 624296 |
| CD43 | BV605 | 1G10 | BD | 563378 |
| Viability Dye | Live dead Aqua | n/a | Invitrogen | L34957 |
| Intracellular | | | | |
| CPT1a | PECy5.5 | ab128568 | Abcam | 8F6AE9 |
| TBET | Pac Blue | 644808 | Biolegend | 644807 |
| GLUT1 | Alexa647 | ab195020 | Abcam | EPR3915 |
| Probes | | | | |
| Streptavidin | BV421 | n/a | Biolegend | 405226 |
| Streptavidin | BV650 | n/a | Biolegend | 405229 |
| Streptavidin | FITC | n/a | Biolegend | 405201 |
| Streptavidin | PE | n/a | Biolegend | 100 |

**Table S5. Innate cell flow cytometry panel configuration**

| Antibody | Fluorophore | Clone | Manufacturer | Catalog # |
|---|---|---|---|---|
| **Surface** | | | | |
| CD11c | BV480 | B-ly6 | Biolegend | 566135 |
| CD123 | APC | 6H6 | Biolegend | 306012 |
| CD25 | BV510 | M-A251 | Biolegend | 563352 |
| CD33 | BV570 | WM53 | Biolegend | 303417 |
| CCR7 | BV650 | G043H7 | Biolegend | 353234 |
| LOX1 | BV421 | 15C4 | Biolegend | 358610 |
| CD38 | BV711 | HIT2 | BD | 563965 |
| CD19 | APC Cy7 | SJ25C1 | Biolegend | 363010 |
| CD56 | APC Cy7 | 5.1H11 | Biolegend | 362512 |
| HLA-DR | BV750 | L243 | Biolegend | 307672 |
| CD3 | BUV661 | UCHT1 | BD | 612964 |
| CD4 | BUV496 | SK3 | BD | 612936 |
| CD8 | BUV 737 | SK1 | BD | 612754 |
| CD16 | BV785/786 | 3G8 | Biolegend | 302046 |
| CD14 | BUV805 | M5E2 | BD | 612902 |
| CD86 | BV605 | BU63 | Biolegend | 374214 |
| CD45RA | BUV563 | HI100 | BD | 612926 |
| CD15 | BUV395 | H198 | BD | 563872 |
| KLRG1 | TRPE | 2F1 | BD | 565393 |
| PD1 | BUV615 | EH12.1 | BD | 612991 |
| CD21 | PE Cy5 | B-ly4 | BD | 551064 |
| CD138 | PE Cy5.5 | MI15 | Biolegend | 356502 |
| CD27 | Spark NIR | O232 | Biolegend | 302856 |
| IgD | BB790 | IA6-2 | BD | 624296 |
| **Intracellular** | | | | |
| FOXP3 | PacBlue | 206D | Biolegend | 320116 |
| Ki67 | PE-Cy7 | Ki-67 | Biolegend | 350526 |
| CPT1a | AF488 | 8F6AE9 | Abcam | ab171449 |
| HK2 | AF680 | EPR20839 | Abcam | ab228819 |
| VDAC1 | AF532 | n/a | Abcam | ab14734 |
| Tomm20 | AF405 | 20B12AF2 | Abcam | ab210047 |
| GLUT1 | Alexa647 | n/a | Abcam | ab195020 |

### Figures

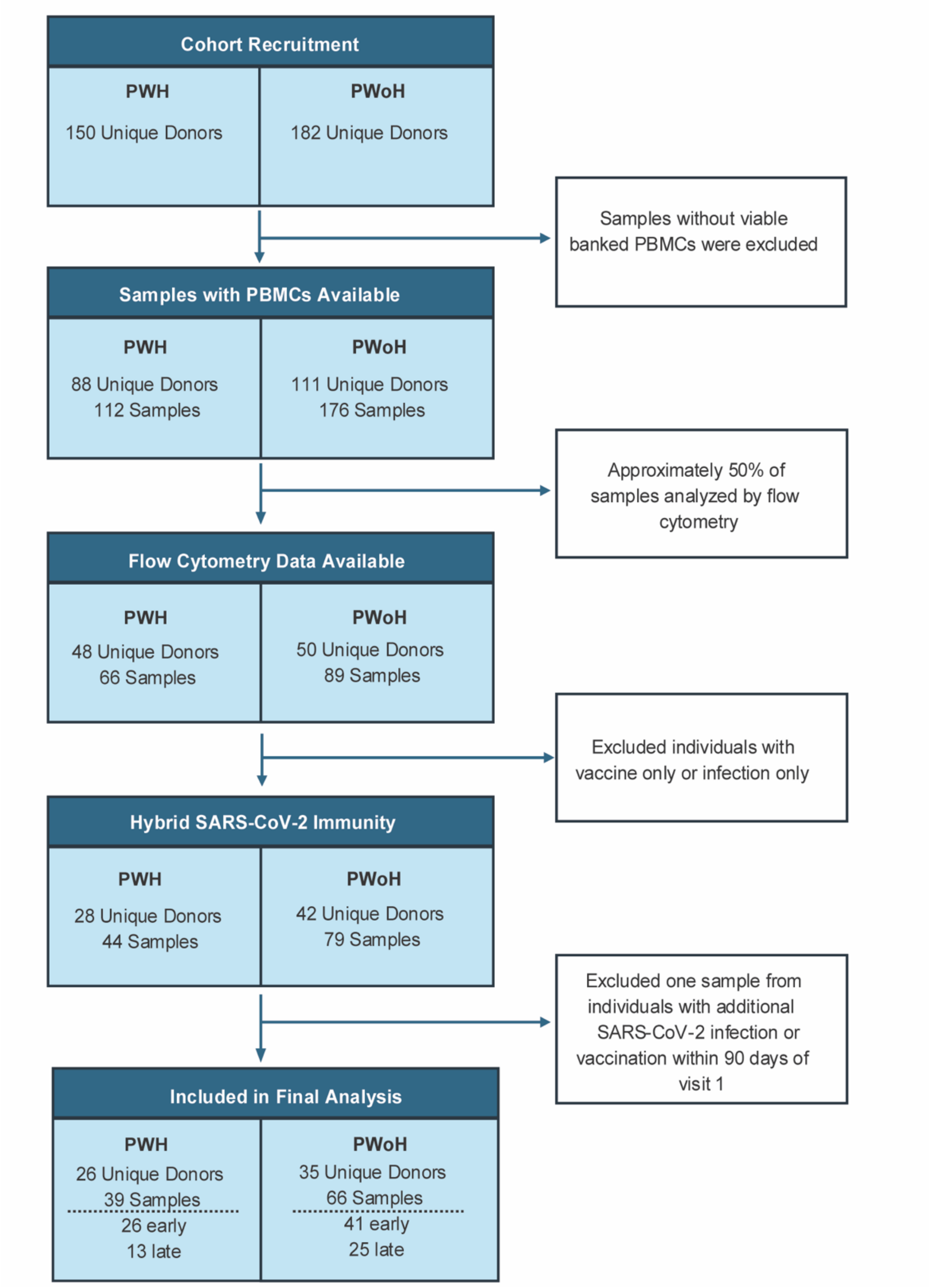


**Figure S1. Flow chart for sample selection.** Inclusion and exclusion criteria for sample selection and final flow cytometry analyses.

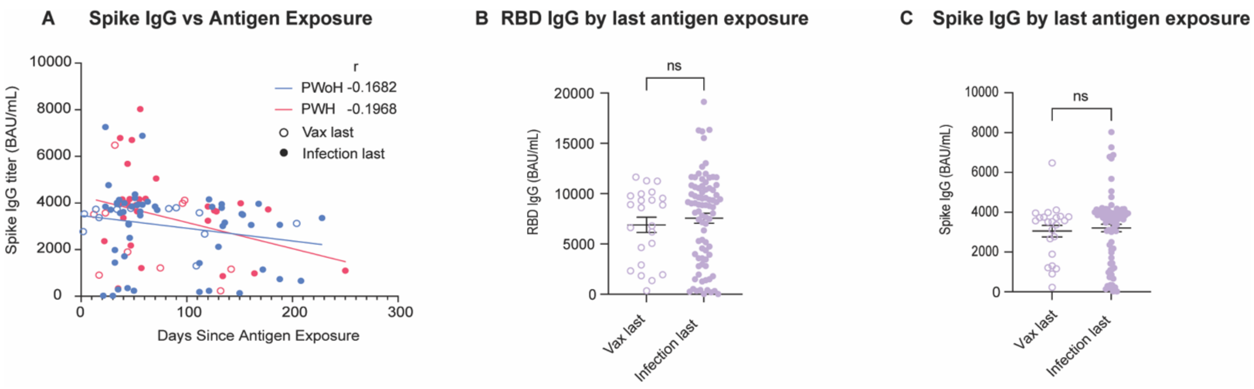


**Figure S2. Anti-spike and anti-RBD antibody responses are correlated among PLWH and HCs, but do not differ between last exposure type. A.** Anti-spike IgG titers are plotted against days since last antigen exposure for PWoH (n=66) and PWH (n=39). Spearman correlation was performed independently for PWoH (blue) and PWH (pink). **B-C.** Anti-RBD IgG titers **(B)** and anti-spike IgG titers **(C)** stratified by last exposure type. Significance was tested using Mann-Whitney U test.

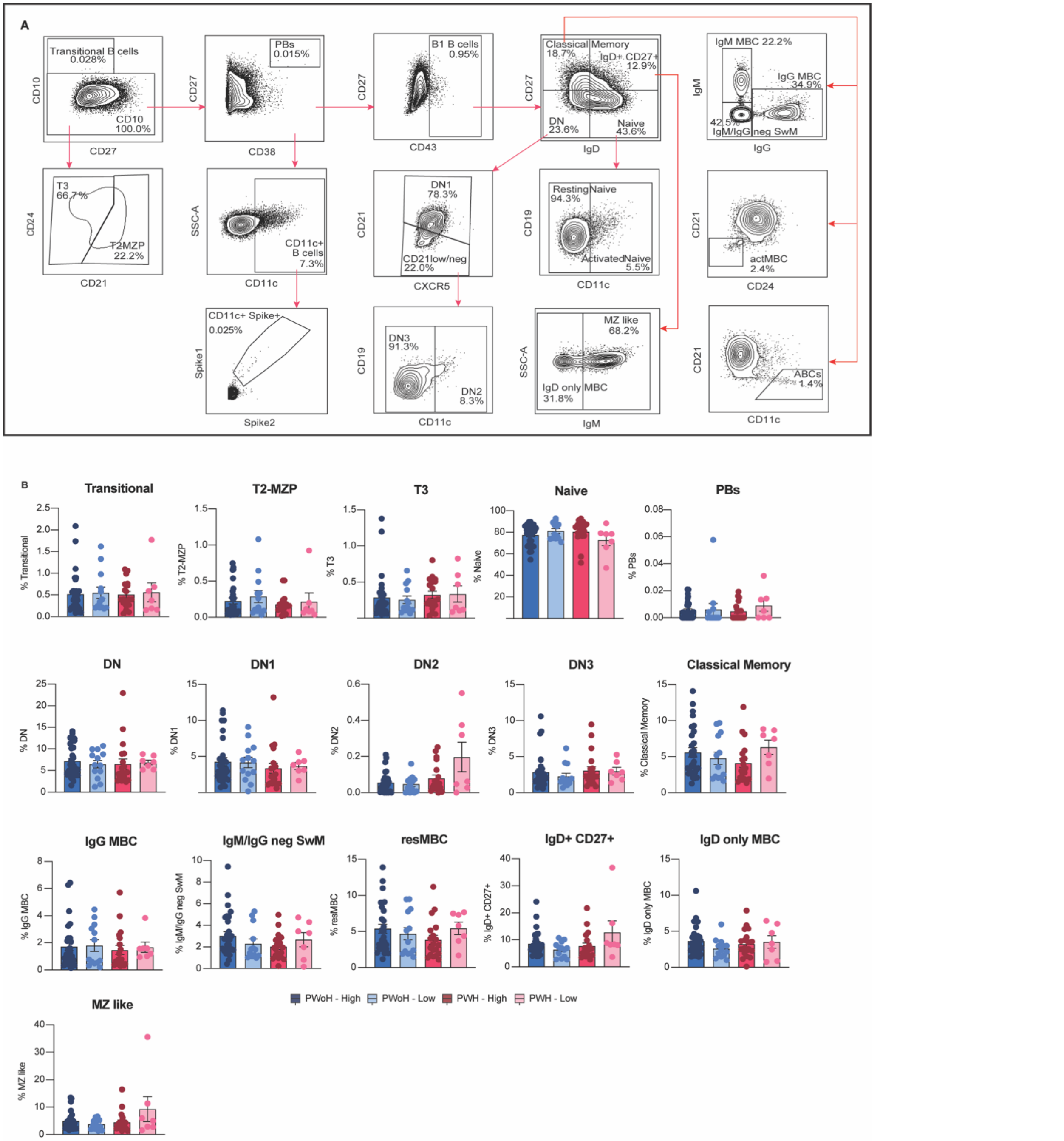

**Figure S3. Frequencies of B cell subsets during the early time point (<90 days since antigen exposure). A.** Representative gating for B cell subsets. **B.** Non-significant B cell subset frequencies (percent of total B cells) stratified by Ab titer and HIV status for the early time point. Significance tested using Kruskal-Wallis test. Each dot represents an individual participant and data are shown as mean±SEM.

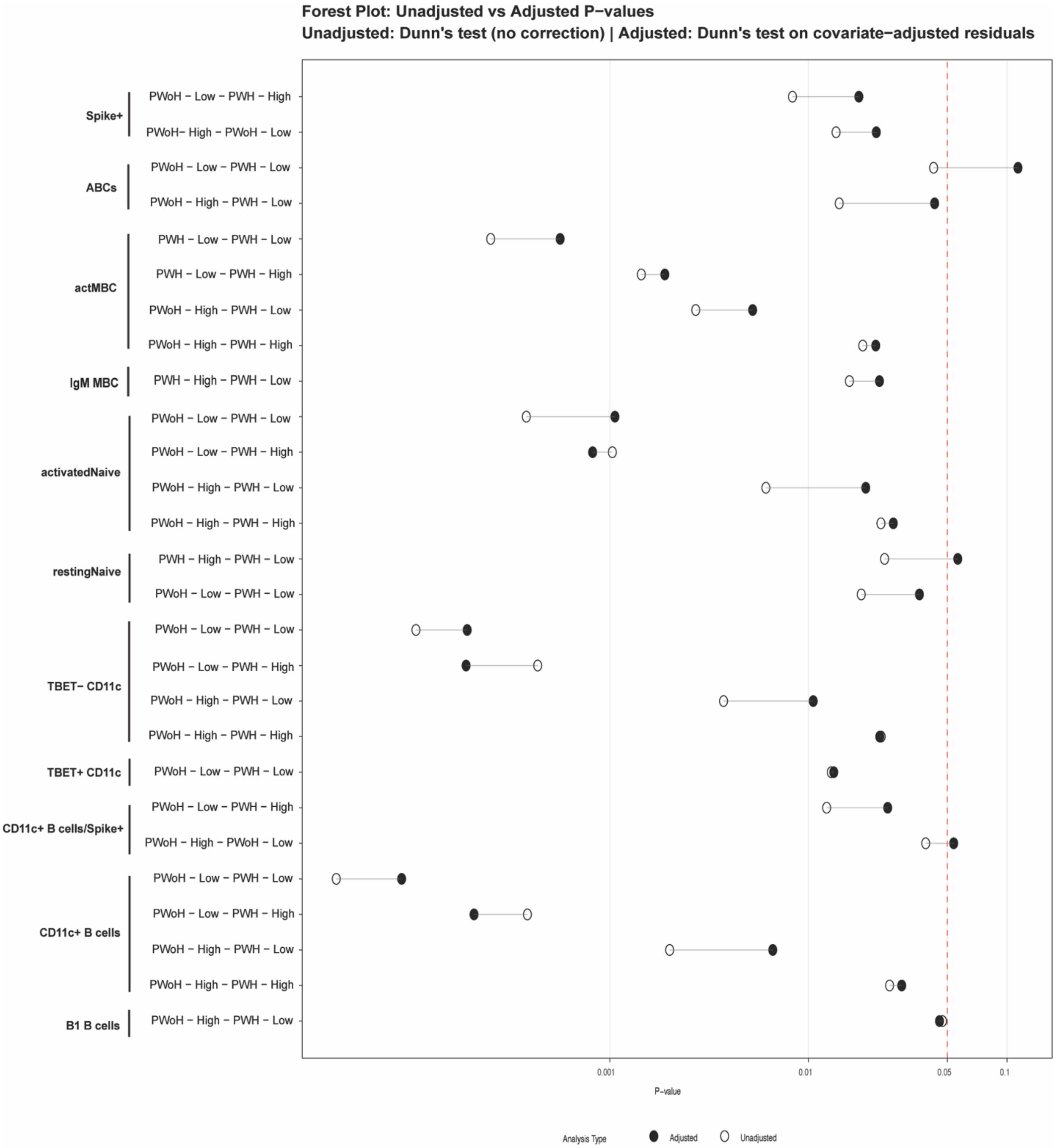


**Figure S4. Adjusted significance values for B cell subset frequencies during the early time point (<90 days since antigen exposure).** Significant B cell subset frequencies identified in Figure 2B-F (open circles) were adjusted for age, BMI, sex, and days since symptom onset (filled circles) using linear regression to calculate covariate-adjusted residuals. All statistical analyses were performed using Kruskal-Wallis test followed by uncorrected Dunn's test. Only comparisons with at least one p-value < 0.5 (unadjusted or adjusted) are shown.

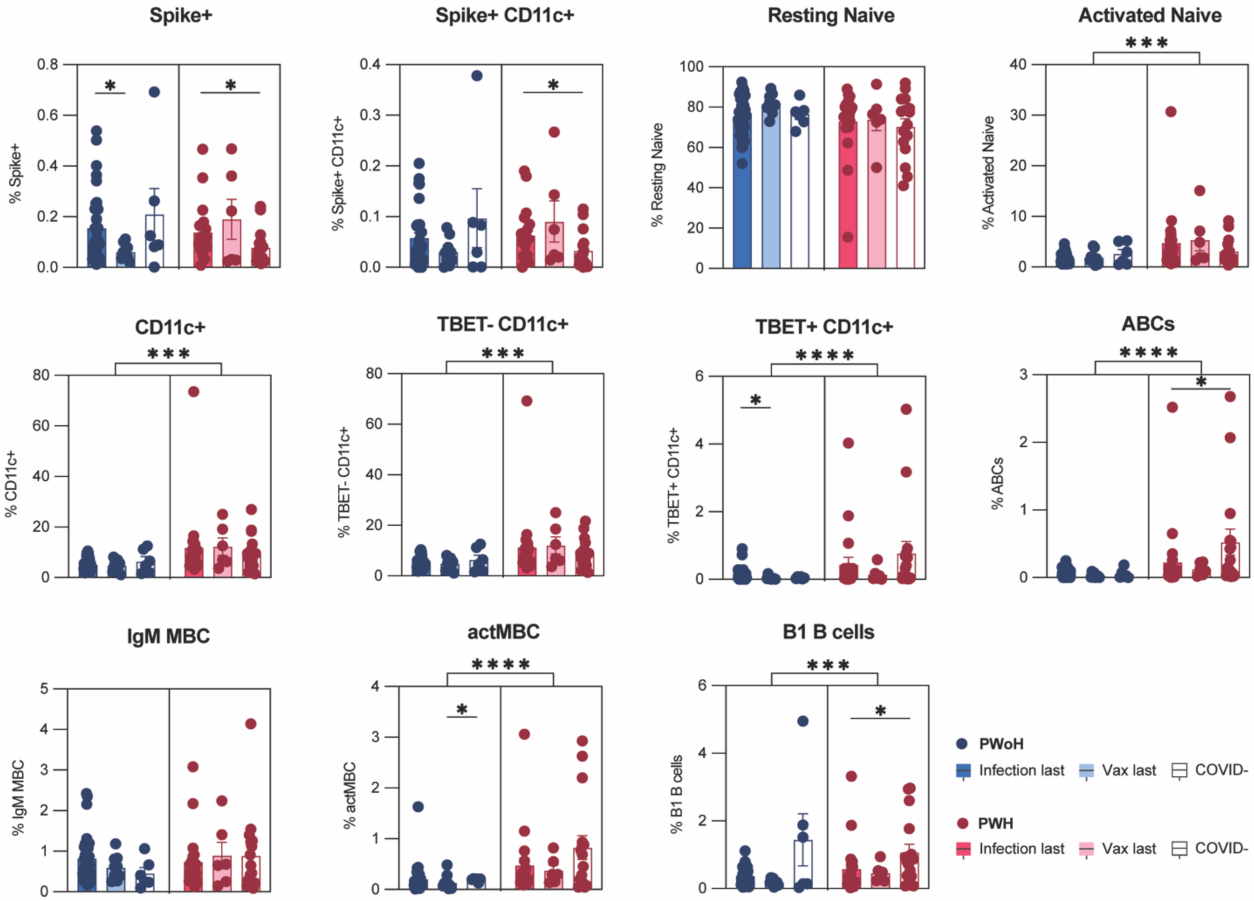


**Figure S5. B cell subset frequencies stratified by last antigen exposure type and HIV status during the early response (<90 days since antigen exposure).** Significant B cell subset frequencies identified in Figure 2B-F were stratified by HIV status and last antigen exposure type for people with hybrid immunity compared to COVID- controls. In-group significance (For PWoH and PWH) tested using Kruskal-Wallis test. Mean frequencies for PWoH vs PWH tested using Mann-Whitney U test. Each dot represents an individual participant and data are shown as mean±SEM. *$p < 0.05$, **$p < 0.01$, ***$p < 0.001$, and ****$p < 0.0001$

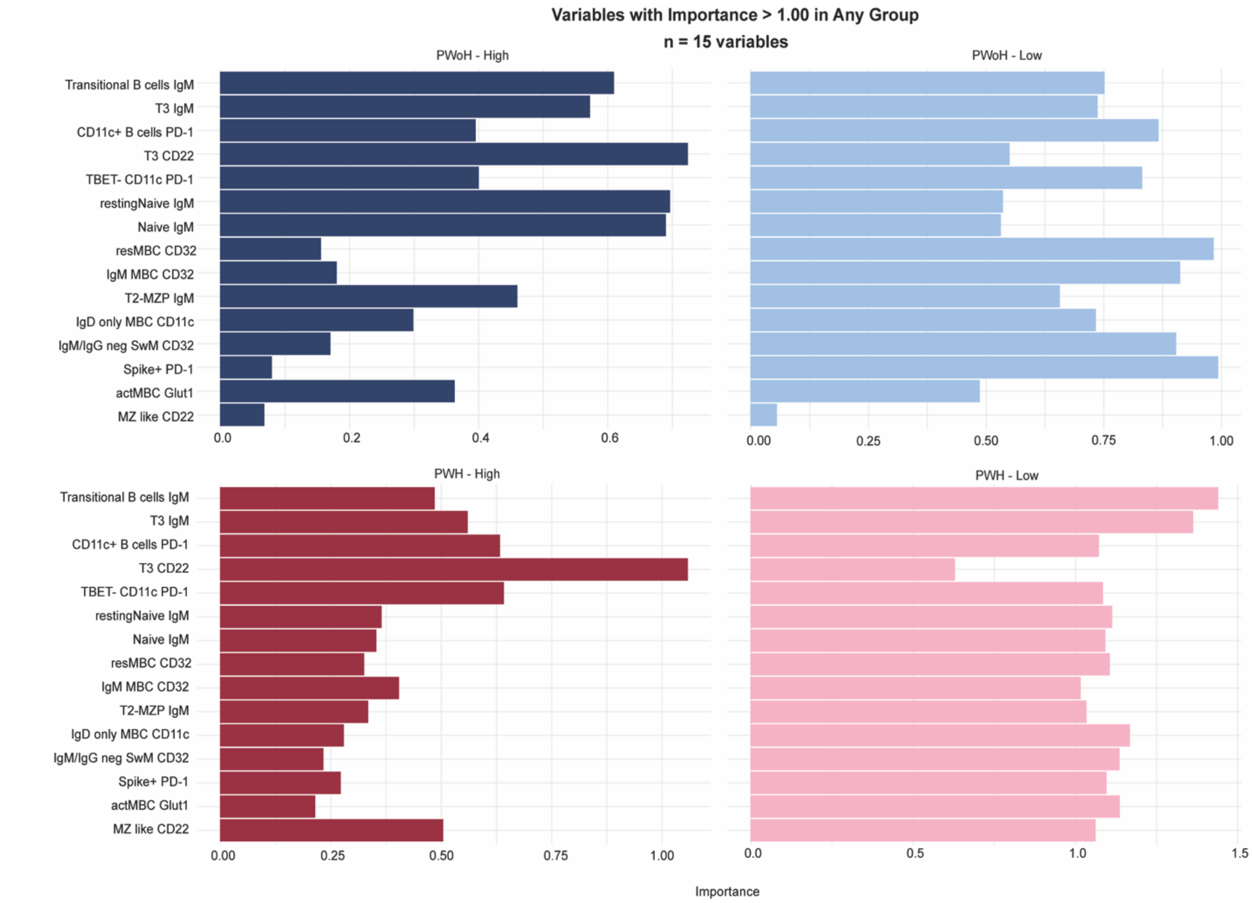


**Figure S6: Top variables for early Ab response group segregation.** Linear discriminant analysis (LDA) followed by effect size filtering was performed to determine top clinical and B cell phenotypic variables for early Ab response group segregation. Variable importance scores (Cohen's d effect size) are shown for any variable with an importance score greater than 1.0 in one or more early Ab response groups (n=15).

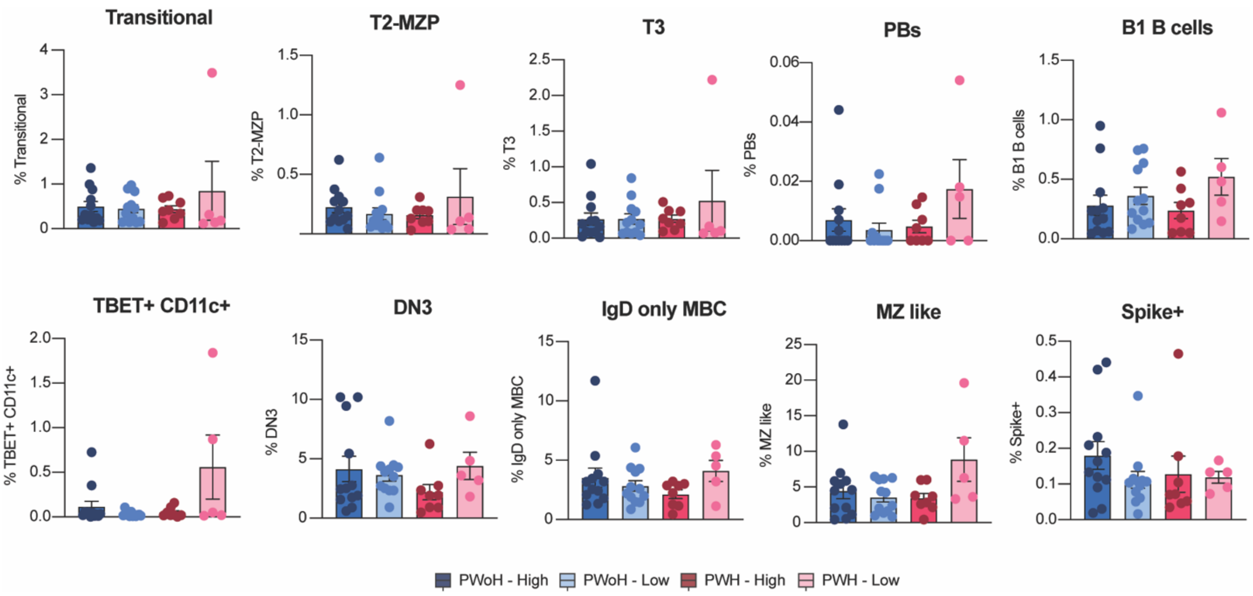


**Figure S7: Frequencies of B cell subsets during the late response (>90 days since antigen exposure).** Non-significant B cell subset frequencies stratified by response type and HIV status for the late response. Significance tested using Kruskal-Wallis test. Each dot represents an individual participant and data are shown as mean±SEM.

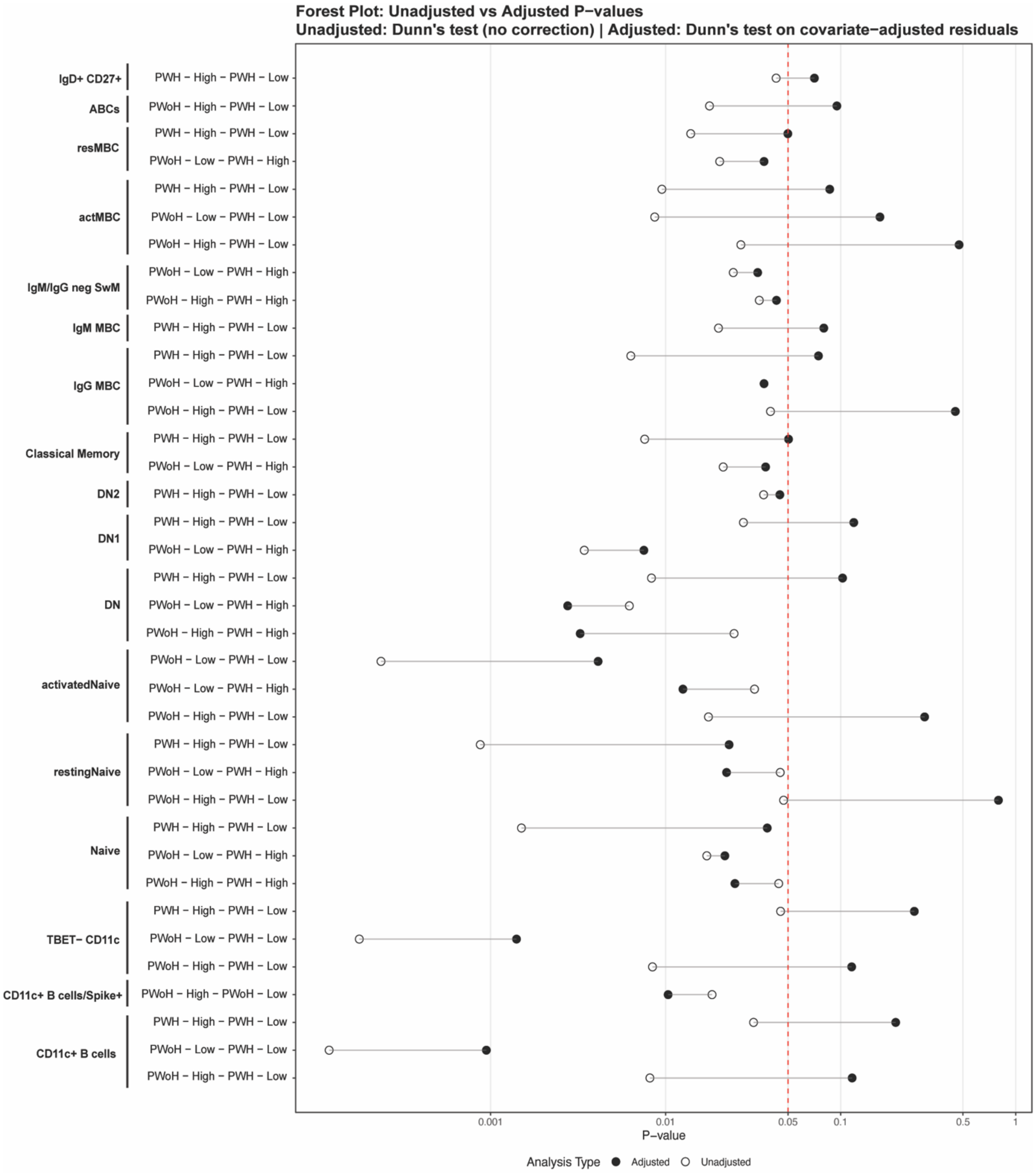


**Figure S8: Adjusted significance values for B cell subset frequencies during the late time point (≥90 days since antigen exposure).** Significant B cell subset frequencies identified in Figure 4B-G (open circles) were adjusted for age, BMI, sex, and days since symptom onset (filled circles) using linear regression to calculate covariate-adjusted residuals. All statistical analyses were performed using Kruskal-Wallis test followed by uncorrected Dunn’s test. Only comparisons with at least one p-value < 0.5 (unadjusted or adjusted) are shown.

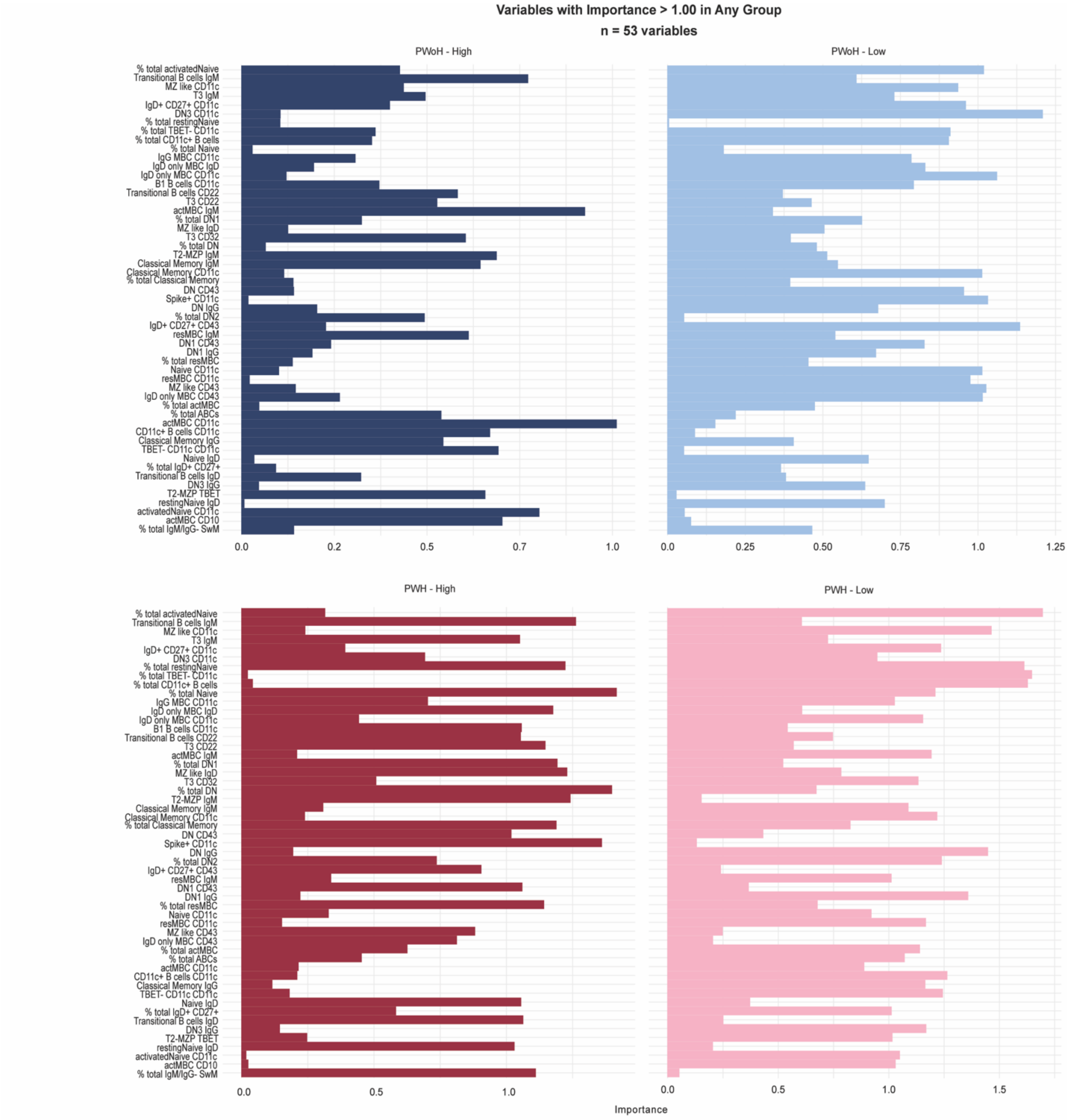


**Figure S9: Top variables for late Ab response group segregation.** Linear discriminant analysis (LDA) followed by effect size filtering was performed to determine top clinical and B cell phenotypic variables for late Ab response group segregation. Variable importance scores (Cohen's d effect size) are shown for any variable with an importance score greater than 1.0 in one or more early Ab response groups (n=53).

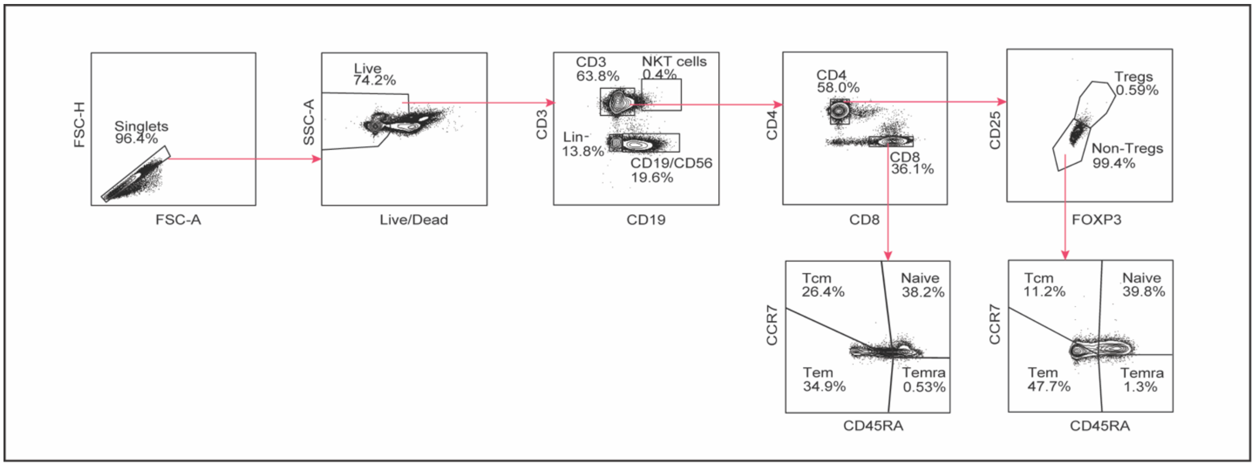


**Figure S10: T cell gating strategy.** Representative gating for T cell subsets.
